# NF-κB Mediated Disruption of Pancreatic Signaling Pathways and its Implications for the Risk of Pancreatic Adenocarcinoma: a systematic review

**DOI:** 10.1101/2024.08.07.24311644

**Authors:** Ovais Shafi, Rahimeen Rajpar, Aakash, Muhammad Waqas, Madiha Haseeb, Raveena, Madhurta Kumari, Ajay Kumar, Muskan Kumari, Muhammad Danial Yaqub

## Abstract

**Objective:** The objective of this study is to determine how NF-κB-mediated inflammation disrupts pancreatic signaling pathways and the resultant increase in the risk of Pancreatic Adenocarcinoma.

**Background:** Pancreatic adenocarcinoma’s severity and poor prognosis underscore the critical role of NF-κB-mediated inflammation in disease progression. NF-κB activation disrupts key pancreatic signaling pathways by promoting cell proliferation, inhibiting differentiation, and facilitating oncogenic transformation. Understanding these mechanisms is essential for developing targeted therapies that mitigate NF-κB-induced damage, improve early detection through biomarkers, and enhance treatment outcomes for pancreatic cancer patients.

**Methods:** Databases, including PubMed, MEDLINE, Google Scholar, and open access/ subscription-based journals were searched for published articles without any date restrictions, to investigate how NF-κB-mediated inflammation disrupts pancreatic signaling pathways and the resultant increase in the risk of Pancreatic Adenocarcinoma. Based on the criteria mentioned in the methods section, studies were systematically reviewed to investigate the research question. This study adheres to relevant PRISMA guidelines (Preferred Reporting Items for Systematic Reviews and Meta-Analyses).

**Results:** NF-κB-mediated inflammation significantly disrupts several critical pancreatic signaling pathways, leading to oncogenic transformations and increased risk of pancreatic adenocarcinoma. Specifically, NF-κB activation alters Notch signaling, causing abnormal cell differentiation; disrupts Wnt and Shh pathways, enhancing cellular proliferation; and perturbs FGF and TGF-β pathways, leading to impaired cellular homeostasis. Additionally, NF-κB interferes with BMP signaling, contributing to dysregulated cell growth; modulates the PI3K/Akt/mTOR pathway, promoting cell survival and tumor growth; affects JAK/STAT signaling, resulting in enhanced inflammatory responses and tumor progression; and disrupts Hippo pathway components, which normally regulate organ size and inhibit tumorigenesis.

**Conclusion:** NF-κB-mediated inflammation disrupts critical pancreas signaling pathways—Notch, Wnt, Hippo, JAK/STAT, and others—promoting cell survival, proliferation, and oncogenic transformation. This dysregulation, seen in chronic inflammation, elevates pancreatic adenocarcinoma risk by fostering a tumorigenic microenvironment. These interactions highlight NF-κB as a central player in pancreatic cancer pathogenesis. Targeting NF-κB and associated pathways emerges as crucial for therapeutic strategies aimed at mitigating inflammatory damage and improving outcomes in pancreatic adenocarcinoma treatment.

## Background

Pancreatic adenocarcinoma is one of the most lethal malignancies, characterized by late diagnosis, limited treatment options, and poor prognosis. Emerging evidence suggests that chronic inflammation plays a key role in the development and progression of pancreatic cancer, with NF-κB signaling pathway activation implicated as a central mechanism in this process. NF-κB is a transcription factor that regulates genes involved in inflammation, immune response, cell survival, and proliferation [1]. In the context of pancreatic cancer, NF-κB activation is often sustained due to factors such as chronic pancreatitis, obesity-related inflammation, or genetic predisposition. This sustained activation leads to dysregulated signaling pathways within the pancreas, profoundly impacting cellular processes crucial for tissue homeostasis and tumorigenesis. NF-κB activation disrupts several key signaling pathways in the pancreas [2, 3]. It induces aberrant Wnt signaling, promoting uncontrolled cell proliferation and inhibiting differentiation, which are critical factors in pancreatic adenocarcinoma development. Inhibition of the Hippo pathway by NF-κB leads to hyperactivation of YAP/TAZ transcription co-activators, promoting cell proliferation, inhibiting apoptosis, and facilitating epithelial-mesenchymal transition (EMT), contributing to tumor progression [4, 5]. Dysregulation of Notch signaling by NF-κB disrupts cell fate determination and differentiation processes, leading to the accumulation of undifferentiated cells prone to oncogenic transformation. Understanding how NF-κB-mediated damage disrupts these signaling pathways is crucial for developing targeted therapies and improving clinical outcomes in pancreatic cancer. Targeting NF-κB and its downstream effectors represents a promising approach to mitigate inflammation-driven pancreatic damage and restore normal signaling pathway activity [6]. Moreover, identifying biomarkers associated with NF-κB activation and dysregulated signaling pathways may improve early detection and personalized treatment strategies for patients at risk of or diagnosed with pancreatic adenocarcinoma. This research study aims to elucidate the molecular mechanisms by which NF-κB damages pancreatic signaling pathways, highlighting its role in pancreatic adenocarcinoma pathogenesis and paving the way for therapeutic interventions to combat this devastating disease [7, 8].

## Methods

### Aim of the Study

The aim of this study is to investigate how NF-κB-mediated inflammation disrupts pancreatic signaling pathways (Notch, Wnt, Shh, FGF, TGF-β, BMP, PI3K/Akt/mTOR, JAK/STAT, and Hippo signaling pathways) and the resultant increase in the risk of Pancreatic Adenocarcinoma. This includes examining the impact of NF-κB dysregulation on these signaling pathways and understanding the mechanisms by which inflammation contributes to pancreatic tumorigenesis. These are also the limitations of the study.

### Research Question

How does NF-κB-mediated inflammation disrupt key pancreatic signaling pathways, leading to an increased risk of Pancreatic Adenocarcinoma?

### Search Focus

A comprehensive literature search was conducted using the PUBMED database, MEDLINE database, and Google Scholar, as well as open access and subscription-based journals. There were no date restrictions for published articles. The search focused on the following key signaling pathways involved in pancreatic signaling, specifically investigating their roles in the context of NF-κB-mediated inflammation and pancreatic adenocarcinoma risk:

- Notch
- Wnt
- Shh (Sonic Hedgehog)
- FGF (Fibroblast Growth Factor)
- TGF-β (Transforming Growth Factor-beta)
- BMP (Bone Morphogenetic Protein)
- PI3K/Akt/mTOR
- JAK/STAT
- Hippo

Screening of the literature was also done on this same basis and related data was extracted. Literature search began in December 2020 and ended in November 2023. An in-depth investigation was conducted during this duration based on the parameters of the study as defined above. During revision, further literature was searched and referenced until March 2024. The literature search and all sections of the manuscript were checked multiple times during the months of revision (April 2024 – July 2024) to maintain the highest accuracy possible. This comprehensive approach ensured that the selected studies provided valuable insights into the inflammatory pathways and their implications in pancreatic adenocarcinoma risk. This study adheres to relevant PRISMA guidelines (Preferred Reporting Items for Systematic Reviews and Meta-Analyses).

### Search Queries/Keywords

1. General Terms:

- “Pancreatic Adenocarcinoma”
- “Pancreatic cancer”
- “NF-κB and inflammation”
- “Pancreatic signaling pathways”
2. Specific Pathways:

- “NF-κB” AND “Notch” AND “Pancreatic Adenocarcinoma”
- “NF-κB” AND “Wnt” AND “Pancreatic Adenocarcinoma”
- “NF-κB” AND “Shh” AND “Pancreatic Adenocarcinoma”
- “NF-κB” AND “FGF” AND “Pancreatic Adenocarcinoma”
- “NF-κB” AND “TGF-β” AND “Pancreatic Adenocarcinoma”
- “NF-κB” AND “BMP” AND “Pancreatic Adenocarcinoma”
- “NF-κB” AND “PI3K/Akt/mTOR” AND “Pancreatic Adenocarcinoma”
- “NF-κB” AND “JAK/STAT” AND “Pancreatic Adenocarcinoma”
- “NF-κB” AND “Hippo” AND “Pancreatic Adenocarcinoma”

Boolean operators (AND, OR) were utilized to construct search queries in relation to the following terms: “Pancreatic Adenocarcinoma,” “NF-κB,” “Inflammation,” and “Signaling Pathways.” These operators were applied in relation to key signaling pathways involved in pancreatic signaling. This approach facilitated the comprehensive retrieval of relevant articles by combining these terms to capture studies focusing on their interrelations and individual contributions to pancreatic tumorigenesis.

### Objectives of the Searches

- To determine the impact of NF-κB-mediated inflammation on disrupting pancreatic signaling pathways.
- To investigate how such disruptions contribute to pancreatic tumorigenesis.
- To assess the role of specific signaling pathways (Notch, Wnt, Shh, FGF, TGF-β, BMP, PI3K/Akt/mTOR, JAK/STAT, and Hippo) in the increased risk of pancreatic adenocarcinoma.

### Screening and Eligibility Criteria

#### Initial Screening

Articles were initially screened based on titles and abstracts to identify direct relevance to the study objectives.

#### Full-Text Review

Articles that passed the initial screening underwent a comprehensive full-text review. Articles were included if they provided valuable insights into the roles of the specified signaling pathways in pancreatic adenocarcinoma in the context of NF-κB-mediated inflammation.

#### Data Extraction

Relevant data was extracted from each selected article, focusing on key findings and outcomes related to the study objectives.

### Inclusion and Exclusion Criteria

#### Inclusion Criteria

- Articles directly related to the key signaling pathways (Notch, Wnt, Shh, FGF, TGF-β, BMP, PI3K/Akt/mTOR, JAK/STAT, Hippo) involved in pancreatic signaling and inflammation.
- Studies focusing on the impact of NF-κB dysregulation on these pathways and pancreatic tumorigenesis.

#### Exclusion Criteria

- Articles that did not conform to the study focus.
- Insufficient methodological rigor.
- Data not aligning with the research questions.

### Rationale for Screening and Inclusion

- **Notch:** Essential for pancreatic development and differentiation. Disruption leads to tumorigenesis.
- **Wnt:** Regulates cell proliferation and differentiation. Dysregulation associated with cancer progression.
- **Shh:** Involved in pancreatic organogenesis. Aberrant signaling linked to tumor growth.
- **FGF:** Important for cell growth and differentiation. Dysregulation contributes to cancer development.
- **TGF-β:** Regulates cell growth and apoptosis. Dysregulation promotes tumor progression.
- **BMP:** Involved in tissue homeostasis. Abnormal signaling linked to cancer.
- **PI3K/Akt/mTOR:** Key pathway in cell survival and proliferation. Dysregulation promotes cancer cell growth.
- **JAK/STAT:** Regulates immune response and cell growth. Aberrant signaling associated with cancer.
- **Hippo:** Controls organ size and cell proliferation. Dysregulation leads to cancer.

This comprehensive screening and inclusion rationale ensure that the selected studies provide valuable insights into the roles of these critical signaling pathways in pancreatic inflammation and tumorigenesis. PRISMA Flow Diagram is Fig 1.

**Fig 1.**
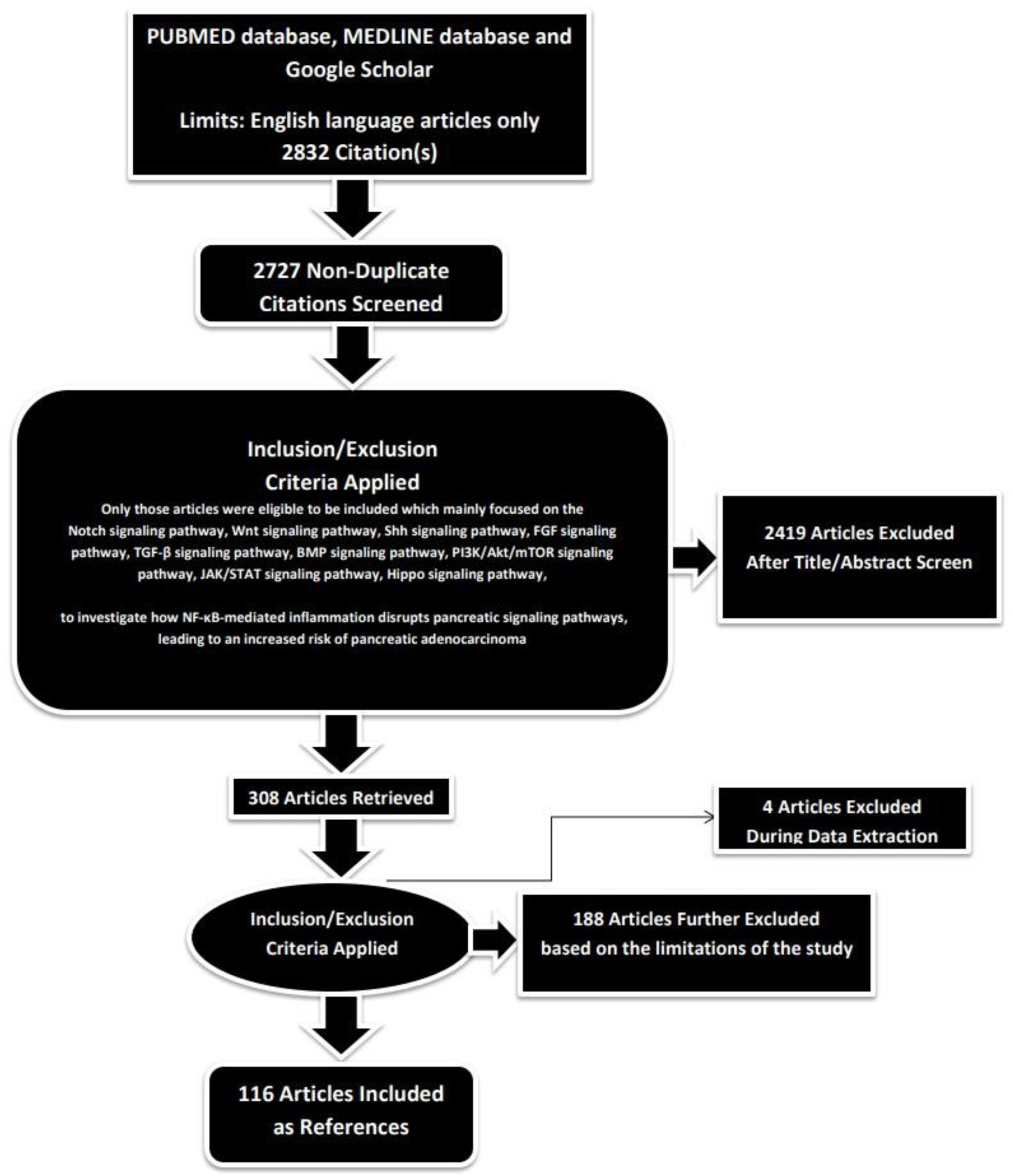
PRISMA FLOW DIAGRAM: This figure represents graphically the flow of citations in the study.

### Assessment of Article Quality and Potential Biases

Ensuring the quality and minimizing potential biases of the selected articles were crucial aspects to guarantee the rigor and reliability of the research findings.

#### Quality Assessment

The initial step in quality assessment involved evaluating the methodological rigor of the selected articles. This included a thorough examination of the study design, data collection methods, and analyses conducted. The significance of the study’s findings was weighed based on the quality of the evidence presented. Articles demonstrating sound methodology— such as well-designed studies, controlled variables, and scientifically robust data—were considered of higher quality. Peer-reviewed articles, scrutinized by experts in the field, served as a significant indicator of quality.

#### Potential Biases Assessment

- **Publication Bias:** To address the potential for publication bias, a comprehensive search strategy was adopted to include a balanced representation of both positive and negative results, incorporating a wide range of published articles from databases like Google Scholar.
- **Selection Bias:** Predefined and transparent inclusion criteria were applied to minimize subjectivity in the selection process. Articles were chosen based on their relevance to the study’s objectives, adhering strictly to these criteria. This approach reduced the risk of subjectivity and ensured that the selection process was objective and consistent.
- **Reporting Bias:** To mitigate reporting bias, articles were checked for inconsistencies or missing data. Multiple detailed reviews of the methodologies and results were conducted for all selected articles to identify and address any reporting bias.

By including high-quality, peer-reviewed studies and thoroughly assessing potential biases, this study aimed to provide a robust foundation for the results and conclusions presented.

### Language and Publication Restrictions

We restricted our selection to publications in the English language. There were no limitations imposed on the date of publication. Unpublished studies were not included in our analysis.

## Results

A total of 2832 articles were identified using database searching, and 2727 were recorded after duplicates removal. 2419 were excluded after screening of title/abstract, 188 were finally excluded, and 4 articles were excluded during data extraction. These exclusions were primarily due to factors such as non-conformity with the study focus, insufficient methodological rigor, or data that did not align with our research question. Finally, 116 articles were included as references.

### Investigating Pancreatic Signaling Pathways

#### 1. Notch Signaling Pathway

##### NF-κB in disrupting this signaling pathway in pancreas

The Notch signaling pathway is integral to pancreas development and differentiation, with dysregulation potentially posing significant implications, particularly in inflammatory contexts [9]. Inflammation, often mediated by NF-κB and its associated cytokines like TNF-α and IL-1β, exerts influence on Notch signaling through several mechanisms. These mechanisms include direct interaction with promoters of Notch pathway genes, thereby altering transcriptional activity [10, 11]. NF-κB can also impact Notch receptors, ligands, and target genes like Hes1, either enhancing or suppressing their expression levels. Inflammatory conditions commonly induce oxidative stress, which further complicates Notch signaling by modifying the stability and function of Notch receptors and ligands through reactive oxygen species (ROS)-mediated protein alterations. Moreover, inflammation triggers epigenetic changes such as DNA methylation and histone modifications, leading to chromatin state modifications of Notch pathway genes, potentially silencing or activating their expression [12]. MicroRNAs (miRNAs) upregulated during inflammation can target and degrade mRNAs encoding Notch pathway components, contributing to their post-transcriptional regulation. Additionally, inflammatory signaling pathways frequently intersect with others like JAK/STAT or MAPK pathways, exerting complex regulatory effects on Notch signaling. NF-κB, in particular, interacts directly with κB sites within Notch pathway gene promoters, influencing their transcriptional activity [13]. This interaction can upregulate Notch ligands such as Jagged1 or Delta-like ligands, thereby enhancing Notch signaling in specific cellular contexts. NF-κB also induces molecules like Numb or Deltex, which inhibit Notch receptor activation, thereby modulating Notch signaling outcomes. In chronic conditions such as chronic pancreatitis, sustained NF-κB activation due to prolonged inflammation perpetuates the upregulation of Notch pathway components. This dysregulation contributes to aberrant cell differentiation and fibrosis within the pancreas, intensifying the inflammatory response and establishing a feedback loop of inflammation and Notch signaling dysregulation [14].

##### Resultant dysregulation in pancreatic cell fate predisposing pancreas to pancreatic adenocarcinoma

The NF-κB pathway and the Notch signaling pathway are crucial in regulating cell fate decisions within the pancreas, where their interaction profoundly influences pancreatic biology. Dysregulation of these pathways, particularly under conditions of chronic inflammation, can significantly disrupt pancreatic cell fate and escalate the risk of developing pancreatic adenocarcinoma [15]. NF-κB activation during chronic inflammation exerts multiple impacts on the Notch signaling pathway, contributing to these processes. NF-κB activation induces aberrant expression of Notch pathway components, including upregulation of Notch ligands like Jagged1 and receptors such as Notch1. This heightened Notch signaling can lead to improper differentiation of pancreatic progenitor cells, impairing their ability to mature into functional exocrine and endocrine cells [16]. Instead, these cells may remain in an undifferentiated or poorly differentiated state, disrupting normal pancreatic architecture and function. Moreover, NF-κB-mediated upregulation of Notch signaling promotes ductal metaplasia, where acinar cells transform into ductal-like cells. This transformation, driven by the inflammatory milieu, serves as a precursor to pancreatic intraepithelial neoplasia (PanIN), a recognized precursor lesion to pancreatic adenocarcinoma [17]. The NF-κB pathway also enhances pancreatic cell proliferation by upregulating Notch target genes that promote cell cycle progression, such as cyclin D1. This heightened proliferation increases the likelihood of acquiring additional genetic mutations, further predisposing cells to malignant transformation. Additionally, NF-κB facilitates cell survival by promoting the expression of anti-apoptotic genes like Bcl-2. Combined with dysregulated Notch signaling, this resistance to apoptosis allows pre-cancerous and cancerous cells to persist and expand. Chronic inflammation, sustained by NF-κB activation, fosters a tumor-permissive microenvironment characterized by inflammatory cytokines (e.g., IL-6, TNF-α) and reactive oxygen species (ROS) that induce DNA damage and genetic instability [18]. This inflammatory milieu, coupled with interactions between NF-κB and Notch signaling in stromal cells such as pancreatic stellate cells, promotes the dense stromal reaction typical of pancreatic adenocarcinoma. This stromal environment supports tumor growth, invasion, and metastasis.

Furthermore, NF-κB and Notch signaling cooperatively induce epithelial-mesenchymal transition (EMT), a critical process enabling cancer cells to acquire invasive properties necessary for metastasis. This transition from epithelial to mesenchymal phenotype facilitates pancreatic cancer cells’ ability to invade surrounding tissues and disseminate to distant organs. Inflammatory conditions driven by NF-κB activation also induce genetic mutations (e.g., KRAS mutations) and epigenetic alterations (e.g., DNA methylation, histone modifications) that activate oncogenes and silence tumor suppressor genes. Dysregulated Notch signaling further contributes to these genetic and epigenetic changes, reinforcing the progression towards pancreatic adenocarcinoma [19].

#### 2. Wnt Signaling Pathway

##### NF-κB in disrupting this signaling pathway in pancreas

The Wnt signaling pathway plays a critical role in regulating cell proliferation, differentiation, and migration during pancreas development [20]. However, inflammation mediated by NF-κB can profoundly impact this pathway, leading to dysregulation that significantly influences pancreatic development and function. Several mechanisms illustrate how inflammation, particularly through NF-κB, may dysregulate gene expression within the Wnt signaling pathway. Inflammatory cytokines like TNF-α and IL-1β activate NF-κB, which in turn can modify the expression of Wnt signaling components. NF-κB influences the transcriptional activity of Wnt ligands, receptors, and target genes by binding directly to their promoters [21]. For example, NF-κB can enhance the expression of Wnt ligands such as Wnt1 or Wnt3a, thereby altering Wnt signaling dynamics. Oxidative stress induced by inflammation can also impact Wnt signaling by altering the stability and function of Wnt proteins and receptors. Reactive oxygen species (ROS) generated during inflammation modify the activity of Wnt signaling components, further contributing to pathway dysregulation. Additionally, inflammatory signals can induce changes in DNA methylation and histone modifications, which affect the chromatin structure of Wnt pathway genes [22]. These epigenetic alterations can lead to either repression or activation of Wnt pathway genes, depending on the specific changes induced by inflammatory mediators. Inflammation-induced microRNAs (miRNAs) can target mRNAs of Wnt pathway components, causing their degradation or translational repression. For instance, certain miRNAs upregulated during inflammation may target Wnt receptors like Frizzled (Fzd) or co-receptors like LRP5/6, thereby inhibiting Wnt signaling. Furthermore, inflammatory signaling pathways often interact with other pathways such as JAK/STAT, MAPK, and PI3K/Akt, resulting in complex regulatory effects on Wnt pathway gene expression. This cross-talk between pathways can further modulate Wnt signaling dynamics in response to inflammatory stimuli. In chronic pancreatitis characterized by sustained NF-κB activation, there is persistent upregulation of Wnt pathway components [23]. This dysregulation contributes to abnormal cell proliferation and fibrosis within the pancreas, exacerbating the inflammatory response and establishing a detrimental cycle of inflammation and dysregulation [24].

##### Resultant dysregulation in pancreatic cell fate predisposing pancreas to pancreatic adenocarcinoma

The NF-κB pathway and the Wnt signaling pathway are essential for maintaining cellular functions and tissue homeostasis, particularly in the pancreas. However, their interaction under inflammatory conditions can lead to significant disruptions in pancreatic cell fate and contribute to the progression of pancreatic adenocarcinoma. NF-κB activation during chronic inflammation plays a key role in impacting the Wnt signaling pathway, thereby exacerbating these disruptions [25]. Chronic inflammation induces NF-κB activation, which in turn upregulates the expression of Wnt ligands (e.g., Wnt1, Wnt3a) and receptors (e.g., Frizzled, LRP5/6). This heightened Wnt signaling enhances cell proliferation within the pancreas, leading to aberrant proliferation of pancreatic cells. Moreover, enhanced Wnt signaling interferes with the differentiation of pancreatic progenitor cells, potentially causing them to remain undifferentiated or differentiate into abnormal cell types, disrupting the typical pancreatic architecture. NF-κB-mediated upregulation of Wnt signaling also contributes to ductal metaplasia, where acinar cells undergo transdifferentiation into ductal-like cells [26]. This process, known as ductal metaplasia, is a precursor to pancreatic intraepithelial neoplasia (PanIN), which can progress to pancreatic adenocarcinoma over time. The interaction between NF-κB and Wnt signaling pathways further predisposes the pancreas to pancreatic adenocarcinoma by promoting cell proliferation and survival. NF-κB enhances the proliferative capacity of pancreatic cells by upregulating Wnt target genes involved in cell cycle progression, such as cyclin D1 and c-Myc [27]. This uncontrolled proliferation increases the accumulation of genetic mutations within pancreatic cells. Additionally, NF-κB facilitates cell survival by upregulating anti-apoptotic genes like Bcl-2, allowing pre-cancerous and cancerous cells to evade apoptosis and survive. The chronic inflammatory microenvironment driven by NF-κB supports tumorigenesis through the production of inflammatory cytokines (e.g., IL-6, TNF-α) and reactive oxygen species (ROS), which induce DNA damage and genetic instability [28]. This environment, coupled with interactions between NF-κB and Wnt signaling in stromal cells such as pancreatic stellate cells, contributes to the dense stromal reaction characteristic of pancreatic adenocarcinoma. This stromal environment promotes tumor growth, invasion, and metastasis. Furthermore, both NF-κB and Wnt signaling pathways can synergistically induce epithelial-mesenchymal transition (EMT), a critical process enabling cancer cells to acquire invasive properties and metastasize to distant organs. The inflammatory milieu created by NF-κB activation also fosters genetic mutations (e.g., KRAS mutations) and epigenetic alterations (e.g., DNA methylation, histone modifications) that activate oncogenes and silence tumor suppressor genes [29]. Dysregulated Wnt signaling further contributes to these genetic and epigenetic changes, amplifying the risk of pancreatic adenocarcinoma development. In chronic pancreatitis, sustained NF-κB activation and inflammation lead to persistent upregulation of Wnt signaling components, promoting cell proliferation, ductal metaplasia, PanIN formation, and ultimately the development of pancreatic adenocarcinoma. This interaction between NF-κB and Wnt signaling pathways exacerbates the inflammatory response, creating a detrimental feedback loop that drives tumor progression. The interplay between NF-κB and Wnt signaling pathways under inflammatory conditions disrupts normal pancreatic cell fate by promoting improper differentiation, ductal metaplasia, and resistance to apoptosis. These disruptions, coupled with enhanced cell proliferation, chronic inflammation, EMT, and genetic/epigenetic alterations, significantly increase the susceptibility of the pancreas to developing pancreatic adenocarcinoma [30].

#### 3. Shh Signaling Pathway

##### NF-κB in disrupting this signaling pathway in pancreas

The Sonic Hedgehog (SHH) signaling pathway is essential for the proper development and differentiation of the pancreas [31]. However, dysregulation of this pathway due to inflammation, particularly mediated by NF-κB, can profoundly impact pancreatic development and function. Inflammatory cytokines such as TNF-α and IL-1β activate NF-κB, enabling it to interact directly with the promoters of SHH pathway genes. This interaction alters the transcriptional activity of SHH components, potentially leading to their upregulation or downregulation, thereby disrupting normal SHH signaling [32]. Moreover, inflammation-induced oxidative stress poses another mechanism through which the SHH signaling pathway may be dysregulated. Reactive oxygen species (ROS) generated during inflammation can modify proteins involved in SHH signaling, thereby altering their stability and function. This oxidative stress can further exacerbate disruptions in SHH pathway activity, influencing pancreatic development and function. Inflammatory signals can also induce epigenetic modifications such as changes in DNA methylation and histone modification patterns [33]. These modifications affect the chromatin state of SHH pathway genes, resulting in either repression or activation of these genes. Additionally, inflammation-induced microRNAs (miRNAs) can target mRNAs of SHH pathway components, leading to their degradation or translational repression, further complicating SHH pathway regulation. Furthermore, inflammatory signaling pathways often cross-talk with other signaling pathways like JAK/STAT, MAPK, and Wnt, which can modulate SHH signaling. This interplay adds complexity to the regulation of SHH pathway genes under inflammatory conditions. NF-κB specifically interacts with SHH pathway genes by binding to κB sites in their promoters, thereby influencing their transcriptional activity. This interaction can either enhance or inhibit the expression of SHH ligands, receptors, and target genes like Gli1. Additionally, NF-κB can induce the expression of inhibitory molecules such as Suppressor of Fused (SUFU), which negatively regulates SHH pathway activity [34]. The transcriptional synergy or antagonism between NF-κB and other critical transcription factors (e.g., GLI) further modulates the transcriptional output of SHH target genes. In chronic pancreatitis, sustained NF-κB activation due to prolonged inflammation can lead to persistent dysregulation of SHH pathway components. This dysregulation contributes to abnormal cell proliferation, fibrosis, and improper differentiation within the pancreas. The interaction between NF-κB and SHH signaling exacerbates the inflammatory response, creating a feedback loop that promotes disease progression. The dysregulation of the SHH signaling pathway due to inflammation, particularly through NF-κB, involves multiple mechanisms including direct promoter interaction, oxidative stress, epigenetic modifications, miRNA regulation, and pathway cross-talk. These interactions disrupt normal pancreatic cell fate by promoting improper differentiation, ductal metaplasia, and resistance to apoptosis. Combined with enhanced cell proliferation, chronic inflammation, epithelial-mesenchymal transition (EMT), and genetic/epigenetic alterations, these disruptions predispose the pancreas to the development of pancreatic adenocarcinoma [35].

##### Resultant dysregulation in pancreatic cell fate predisposing pancreas to pancreatic adenocarcinoma

Chronic inflammation, mediated by NF-κB, significantly disrupts SHH signaling, thereby affecting pancreatic cell fate and increasing susceptibility to cancer [36]. NF-κB activation in response to inflammation upregulates SHH pathway components, intensifying SHH signaling. This heightened signaling promotes excessive cell proliferation and hinders normal differentiation processes, leading to the formation of poorly differentiated cell types that alter the pancreas’s normal structure and function [37]. Furthermore, NF-κB-mediated enhancement of SHH signaling contributes to ductal metaplasia, where acinar cells undergo transdifferentiation into ductal-like cells. This transformation, known as ductal metaplasia, serves as a precursor to PanIN (Pancreatic Intraepithelial Neoplasia), a recognized precursor lesion for pancreatic adenocarcinoma [38]. The chronic inflammatory milieu, coupled with amplified SHH signaling, facilitates the formation and progression of PanINs, which can accumulate additional mutations over time, advancing to invasive pancreatic adenocarcinoma. NF-κB not only promotes heightened cell proliferation through SHH signaling but also enhances cell survival mechanisms [39]. By upregulating SHH target genes like Cyclin D1, Myc, and anti-apoptotic gene Bcl-2, NF-κB supports uncontrolled cell proliferation and suppresses apoptosis. This dual effect sustains the survival and expansion of pre-cancerous and cancerous cells within the pancreas. Moreover, NF-κB-driven chronic inflammation establishes a tumor-supportive microenvironment conducive to tumorigenesis. Inflammatory cytokines such as IL-6 and TNF-α, along with reactive oxygen species (ROS), induce DNA damage and genetic instability, further predisposing cells to malignant transformation [40]. The inflammatory and SHH signaling activities in stromal cells like pancreatic stellate cells enhance the desmoplastic reaction, contributing to the dense stromal environment characteristic of pancreatic adenocarcinoma [41]. This supportive stroma fosters tumor growth, invasion, and metastasis. Both NF-κB and SHH signaling pathways can induce Epithelial-Mesenchymal Transition (EMT), where epithelial cells acquire mesenchymal traits that facilitate metastasis. This phenotypic change is crucial for pancreatic cancer cells to invade surrounding tissues and disseminate to distant organs, thereby exacerbating disease progression. Furthermore, the inflammatory environment driven by NF-κB induces genetic mutations (e.g., KRAS) and epigenetic changes (e.g., DNA methylation, histone modifications), complemented by dysregulated SHH signaling, which further drives tumorigenesis. These alterations activate oncogenes and suppress tumor suppressor genes, perpetuating the malignant transformation process [42].

#### 4. FGF Signaling Pathway

##### NF-κB in disrupting this signaling pathway in pancreas

The Fibroblast Growth Factor (FGF) signaling pathway is crucial for pancreas development, influencing the proliferation and differentiation of pancreatic progenitor cells [43]. However, inflammation, particularly mediated by NF-κB, can lead to dysregulation of FGF signaling, thereby disrupting pancreatic development and function. Inflammatory cytokines like TNF-α and IL-1β activate NF-κB, which in turn affects the expression of FGF ligands, receptors, and downstream signaling components [44]. NF-κB can directly bind to κB sites within the promoters of FGF pathway genes, altering their transcriptional activity and potentially causing either upregulation or downregulation of FGF pathway components. Additionally, inflammation induces oxidative stress, which further complicates FGF signaling by modifying the activity and stability of its components. Reactive oxygen species (ROS) generated during inflammation can interfere with FGF ligand binding, receptor activation, or downstream signaling cascades, thereby disrupting the normal functioning of the pathway [45]. Epigenetic modifications triggered by inflammation, such as changes in DNA methylation and histone modifications, also play a role in FGF signaling dysregulation. These modifications can alter the accessibility of FGF pathway gene promoters, influencing their transcriptional activity under inflammatory conditions and potentially leading to aberrant expression patterns of FGF pathway genes [46]. Furthermore, inflammatory signals can induce specific miRNAs that target FGF pathway mRNAs, leading to their degradation or translational repression. This regulation can directly impact FGF receptor activity or downstream signaling molecules, further contributing to the dysregulation of FGF signaling observed in inflamed pancreatic tissue. NF-κB activation in response to inflammation also initiates cross-talk with other signaling pathways such as MAPK and PI3K/Akt, which interact with FGF signaling. This interplay can modulate the activity and expression of FGF pathway components, influencing pancreatic cell fate and function in complex ways. For instance, NF-κB can enhance the expression of certain FGF ligands or receptors through direct promoter binding, thereby promoting FGF signaling and its downstream effects. Conversely, NF-κB activation may also induce the expression of inhibitory molecules like Sprouty proteins, which negatively regulate FGF receptor activation and downstream signaling, contributing to the dysregulation of FGF signaling under inflammatory conditions. In chronic pancreatitis, prolonged NF-κB activation due to persistent inflammation can sustain dysregulation of the FGF signaling pathway. This disruption interferes with the balance between cell proliferation and differentiation in pancreatic progenitor cells, potentially leading to pancreatic fibrosis and the formation of pre-cancerous lesions [47]. This scenario underscores the critical role of NF-κB-mediated inflammation in perturbing FGF signaling and its implications for pancreatic health and disease progression [48].

##### Resultant dysregulation in pancreatic cell fate predisposing pancreas to pancreatic adenocarcinoma

The NF-κB pathway exerts a profound influence on the Fibroblast Growth Factor (FGF) signaling pathway, thereby significantly disrupting pancreatic cell fate and predisposing the pancreas to pancreatic adenocarcinoma. Chronic inflammation activates NF-κB, which in turn upregulates FGF ligands and receptors [49]. This heightened FGF signaling promotes excessive cell proliferation and disrupts the delicate balance between proliferation and differentiation in pancreatic progenitor cells. Consequently, dysregulated FGF signaling, under the influence of NF-κB, can lead to improper differentiation of pancreatic progenitor cells, potentially resulting in either undifferentiated cells or abnormal differentiation patterns that contribute to pancreatic dysfunction [50]. Moreover, NF-κB-mediated dysregulation of the FGF signaling pathway plays a critical role in the formation of pre-cancerous lesions such as Pancreatic Intraepithelial Neoplasia (PanIN). Here, dysregulated FGF signaling contributes to abnormal proliferation and differentiation of pancreatic epithelial cells, paving the way for the progression to pancreatic adenocarcinoma. NF-κB activation also enhances the expression of FGF pathway components, including FGF ligands (e.g., FGF1, FGF2) and receptors (e.g., FGFR1, FGFR2), thereby sustaining proliferative signaling that drives the expansion of pancreatic epithelial cells [51]. This proliferative advantage, coupled with NF-κB-induced upregulation of anti-apoptotic genes, confers a survival advantage to pre-cancerous cells, enabling them to resist apoptosis and accumulate additional mutations that promote carcinogenesis. Furthermore, the inflammatory microenvironment orchestrated by NF-κB promotes a pro-tumorigenic milieu within the pancreas. Inflammatory cytokines (e.g., IL-6, TNF-α) and reactive oxygen species (ROS) generated during chronic inflammation contribute to genetic instability and facilitate the progression of pre-cancerous lesions to invasive pancreatic adenocarcinoma. Additionally, dysregulated FGF signaling, exacerbated by chronic inflammation, enhances stromal interaction and fibrosis, fostering a desmoplastic reaction that supports tumor growth, invasion, and metastasis [52]. NF-κB activation and dysregulated FGF signaling can also induce epithelial-mesenchymal transition (EMT), a critical process that enhances cancer cells’ migratory and invasive capabilities, thereby facilitating metastasis to distant organs. Moreover, the inflammatory environment induced by NF-κB promotes genetic mutations, including common mutations like KRAS mutations, which are key in driving the initiation and progression of pancreatic adenocarcinoma. Dysregulated FGF signaling further contributes to these genetic alterations, amplifying the mutational burden that fuels tumor development [53]. In the context of chronic pancreatitis, sustained NF-κB activation perpetuates dysregulation of the FGF signaling pathway, disrupting pancreatic cell fate by fostering abnormal proliferation, impairing differentiation, and promoting the development of PanIN lesions. This cascade of events underscores how ongoing NF-κB-mediated inflammation and dysregulated FGF signaling collaborate to drive the progression from precancerous changes to invasive pancreatic adenocarcinoma, highlighting the critical interplay between these pathways in pancreatic cancer development [54].

#### 5. TGF Beta Signaling Pathway

##### NF-κB in disrupting this signaling pathway in pancreas

The Transforming Growth Factor-beta (TGF-β) signaling pathway plays a crucial role in pancreas development by regulating cell proliferation, differentiation, and tissue homeostasis [55]. Dysregulation of this pathway, particularly due to inflammation mediated by NF-κB, can disrupt pancreatic development and potentially contribute to pancreatic diseases. NF-κB-mediated inflammation exerts its influence on the TGF-β pathway through several mechanisms. Activation of NF-κB by inflammatory cytokines such as TNF-α and IL-1β can modulate TGF-β pathway components at various levels. NF-κB can directly bind to κB sites in the promoters of TGF-β pathway genes, thereby altering their transcriptional activity [56]. This regulatory process can lead to either the upregulation or downregulation of TGF-β ligands, receptors, and downstream signaling molecules, influencing cellular responses related to growth and differentiation. Inflammatory conditions also induce oxidative stress, which further complicates TGF-β signaling. Reactive oxygen species (ROS) generated under inflammatory settings can modify TGF-β receptors or downstream effectors, potentially altering their signaling capabilities and perturbing cellular responses mediated by TGF-β. Additionally, inflammation can induce epigenetic modifications such as changes in DNA methylation and histone modifications [57]. These alterations affect the accessibility of TGF-β pathway gene promoters, thereby impacting the transcriptional activity of TGF-β ligands, receptors, and intracellular signaling mediators. Such epigenetic changes under inflammatory conditions can significantly influence cellular processes governed by TGF-β signaling. Furthermore, inflammatory signals can induce specific microRNAs (miRNAs) that target TGF-β pathway mRNAs, leading to their degradation or translational repression [58]. Certain miRNAs upregulated during inflammation may directly inhibit TGF-β receptors or downstream signaling molecules, adding another layer of complexity to the dysregulation of TGF-β signaling in pancreatic cells. NF-κB activation not only interacts directly with TGF-β pathway gene promoters but also engages in cross-talk with other signaling pathways such as MAPK and PI3K/Akt. This cross-talk can modulate the activity and expression of TGF-β pathway components, thereby influencing pancreatic cell fate and function in various contexts, including development and disease progression. In chronic pancreatitis, sustained NF-κB activation due to persistent inflammation disrupts the TGF-β signaling pathway over time [59]. This dysregulation impairs the balance between cell proliferation, differentiation, and apoptosis in pancreatic cells, potentially contributing to fibrosis and the development of pancreatic diseases, including pancreatic adenocarcinoma. The interplay between NF-κB-mediated inflammation and TGF-β signaling highlights the mechanisms by which inflammatory processes can impact pancreatic cellular dynamics and disease pathogenesis [60].

##### Resultant dysregulation in pancreatic cell fate predisposing pancreas to pancreatic adenocarcinoma

The Transforming Growth Factor-beta (TGF-β) signaling pathway plays a crucial role in pancreas development by regulating cell proliferation, differentiation, and tissue homeostasis [61]. Dysregulation of this pathway, particularly due to inflammation mediated by NF-κB, can disrupt pancreatic development and potentially contribute to pancreatic diseases. NF-κB-mediated inflammation exerts its influence on the TGF-β pathway through several mechanisms. Activation of NF-κB by inflammatory cytokines such as TNF-α and IL-1β can modulate TGF-β pathway components at various levels [62]. NF-κB can directly bind to κB sites in the promoters of TGF-β pathway genes, thereby altering their transcriptional activity. This regulatory process can lead to either the upregulation or downregulation of TGF-β ligands, receptors, and downstream signaling molecules, influencing cellular responses related to growth and differentiation. Inflammatory conditions also induce oxidative stress, which further complicates TGF-β signaling. Reactive oxygen species (ROS) generated under inflammatory settings can modify TGF-β receptors or downstream effectors, potentially altering their signaling capabilities and perturbing cellular responses mediated by TGF-β [63]. Additionally, inflammation can induce epigenetic modifications such as changes in DNA methylation and histone modifications. These alterations affect the accessibility of TGF-β pathway gene promoters, thereby impacting the transcriptional activity of TGF-β ligands, receptors, and intracellular signaling mediators. Such epigenetic changes under inflammatory conditions can significantly influence cellular processes governed by TGF-β signaling. Furthermore, inflammatory signals can induce specific microRNAs (miRNAs) that target TGF-β pathway mRNAs, leading to their degradation or translational repression [64]. Certain miRNAs upregulated during inflammation may directly inhibit TGF-β receptors or downstream signaling molecules, adding another layer of complexity to the dysregulation of TGF-β signaling in pancreatic cells. NF-κB activation not only interacts directly with TGF-β pathway gene promoters but also engages in cross-talk with other signaling pathways such as MAPK and PI3K/Akt. This cross-talk can modulate the activity and expression of TGF-β pathway components, thereby influencing pancreatic cell fate and function in various contexts, including development and disease progression. In chronic pancreatitis, sustained NF-κB activation due to persistent inflammation disrupts the TGF-β signaling pathway over time [65]. This dysregulation impairs the balance between cell proliferation, differentiation, and apoptosis in pancreatic cells, potentially contributing to fibrosis and the development of pancreatic diseases, including pancreatic adenocarcinoma. The interplay between NF-κB-mediated inflammation and TGF-β signaling highlights the mechanisms by which inflammatory processes can impact pancreatic cellular dynamics and disease pathogenesis [66, 67].

#### 6. BMP Signaling Pathway

##### NF-κB in disrupting this signaling pathway in pancreas

The Bone Morphogenetic Protein (BMP) signaling pathway is essential for pancreas development, influencing critical processes such as pancreatic progenitor specification, differentiation, and the maintenance of pancreatic cell identity. Dysregulation of BMP signaling, particularly through inflammation mediated by NF-κB, can disrupt pancreatic development and potentially contribute to pancreatic diseases [68]. NF-κB-mediated inflammation impacts the BMP signaling pathway through several interconnected mechanisms. Inflammatory cytokines like TNF-α and IL-1β activate NF-κB, enabling direct influence over BMP pathway components. NF-κB can bind to κB sites in the promoters of BMP pathway genes, thereby modulating their transcriptional activity [69]. This regulatory role can result in either the enhancement or reduction of BMP ligands (such as BMP2, BMP4, BMP7), receptors (including BMPR1A, BMPR1B, BMPR2), and downstream signaling molecules like Smads, crucial for transmitting BMP signals within cells. Inflammatory conditions also induce oxidative stress, which further complicates BMP signaling. Reactive oxygen species (ROS) generated under inflammatory stress can alter BMP receptor activation or downstream signaling cascades, potentially impairing BMP pathway activity and disrupting its normal cellular responses in pancreatic development and maintenance [70]. Moreover, inflammation-induced epigenetic modifications, such as changes in DNA methylation patterns and histone modifications, influence the accessibility of BMP pathway gene promoters. These alterations in epigenetic regulation may lead to dysregulated expression of BMP ligands and receptors under inflammatory conditions, further complicating the finely tuned balance required for proper BMP signaling. Inflammatory signals can also induce specific microRNAs (miRNAs) that target BMP pathway mRNAs, resulting in their degradation or translational repression [71]. Elevated levels of certain miRNAs during inflammation may directly inhibit BMP receptors or downstream signaling molecules, thereby altering BMP pathway activity and disrupting its role in pancreatic cell fate determination and function. NF-κB activation not only directly impacts BMP pathway gene transcription but also interacts with other signaling pathways like MAPK and PI3K/Akt, forming a complex network of signaling cross-talk [72]. This interplay can modulate BMP pathway activity and gene expression, influencing pancreatic cell fate decisions, differentiation processes, and responses to pathological conditions. In chronic pancreatitis, sustained NF-κB activation due to ongoing inflammation can dysregulate BMP signaling pathways. This dysregulation may lead to excessive fibrosis by aberrantly activating pancreatic stellate cells through BMP-mediated mechanisms. Furthermore, dysregulated BMP signaling can disrupt normal pancreatic progenitor cell differentiation, potentially promoting the formation of abnormal cell populations that contribute to the development of pancreatic diseases, including pancreatic cancer [73].

##### Resultant dysregulation in pancreatic cell fate predisposing pancreas to pancreatic adenocarcinoma

The impact of NF-κB on the Bone Morphogenetic Protein (BMP) signaling pathway is profound, contributing to disruptions in pancreatic cell fate and potentially predisposing the pancreas to pancreatic adenocarcinoma through several interconnected mechanisms [74]. Chronic inflammation triggers sustained NF-κB activation, which in turn dysregulates BMP signaling by influencing the expression of BMP ligands (such as BMP2, BMP4, BMP7), receptors (including BMPR1A, BMPR1B, BMPR2), and downstream signaling molecules like Smads. This dysregulation impedes the differentiation of pancreatic progenitor cells, crucial for the formation of mature pancreatic cell types—both endocrine and exocrine. Such impairment not only compromises pancreatic function but also heightens susceptibility to various pancreatic diseases. NF-κB-mediated dysregulation of BMP signaling also plays a key role in promoting fibrosis within the pancreas. By altering BMP signaling pathways, NF-κB induces a fibrogenic response that leads to excessive fibrosis, thereby restructuring pancreatic tissue architecture. This fibrotic environment not only impairs organ function but also creates a favorable microenvironment conducive to tumor growth, particularly observed in pancreatic adenocarcinoma [75]. Furthermore, NF-κB activation exacerbates stromal activation, notably through pancreatic stellate cells, which further contributes to the desmoplastic reaction characteristic of pancreatic cancer. In terms of cancer predisposition, NF-κB activation enhances BMP signaling activity, thereby promoting cell proliferation and survival pathways in pancreatic cells [76]. This enhancement supports the expansion of pre-cancerous lesions and facilitates tumor progression. Dysregulated BMP signaling, influenced by NF-κB, also inhibits apoptosis by upregulating anti-apoptotic proteins like Bcl-2, which enhances the survival of cancerous cells and sustains tumor growth [77]. Moreover, NF-κB-mediated dysregulation of BMP signaling can induce epithelial-mesenchymal transition (EMT) in pancreatic epithelial cells [78]. EMT is a critical process in cancer metastasis, enabling cancer cells to acquire migratory and invasive properties necessary for metastasis to distant organs. Additionally, dysregulated BMP signaling may suppress the immune response within the tumor microenvironment, facilitating immune evasion by cancer cells and perpetuating a pro-inflammatory milieu that supports tumor growth and progression [79, 80].

#### 7. PI3K/Akt/mTOR Signaling Pathway

##### NF-κB in disrupting this signaling pathway in pancreas

The PI3K/Akt/mTOR signaling pathway plays a critical role in pancreas development by regulating essential processes such as cell growth, survival, metabolism, and differentiation [81]. However, dysregulation of this pathway due to inflammation, particularly mediated by NF-κB, can disrupt pancreatic homeostasis and contribute to various pancreatic diseases. Several potential mechanisms illustrate how NF-κB-mediated inflammation may influence the gene expression and activity of the PI3K/Akt/mTOR signaling pathway. Inflammatory cytokines like TNF-α and IL-1β activate NF-κB, enabling it to directly impact components of the PI3K/Akt/mTOR pathway [82]. NF-κB can bind to κB sites within the promoters of genes encoding PI3K isoforms (such as PI3Kα, PI3Kβ), Akt isoforms (including Akt1, Akt2, Akt3), mTOR, and downstream effectors like S6K and 4E-BP. This binding modulates their transcriptional activity, leading to either the upregulation or downregulation of these crucial pathway components, thereby influencing pathway activity [83]. Additionally, inflammatory conditions induce oxidative stress, which further complicates PI3K/Akt/mTOR pathway regulation. Reactive oxygen species (ROS) generated during inflammation can alter the phosphorylation status of Akt and mTOR, directly impacting their signaling activities. Moreover, inflammation-associated epigenetic modifications, including changes in DNA methylation patterns and histone modifications, can affect the accessibility of PI3K/Akt/mTOR pathway gene promoters. These alterations in epigenetic regulation can consequently influence the expression levels of PI3K isoforms, Akt isoforms, and mTOR, thereby affecting overall pathway activity [84]. Furthermore, inflammatory signals can induce specific microRNAs (miRNAs) that target mRNAs encoding PI3K/Akt/mTOR pathway components, leading to their degradation or translational repression. Certain miRNAs upregulated during inflammation may directly inhibit PI3K isoforms, Akt isoforms, or mTOR, thereby modulating pathway activity in response to inflammatory stimuli. NF-κB activation also facilitates cross-talk with other signaling pathways like MAPK and TGF-β, which interact with PI3K/Akt/mTOR signaling. This interplay can further modulate the activity and expression of pathway components, influencing pancreatic cell fate, growth, and metabolic functions in complex ways. An example illustrating this dysregulation is seen in the context of pancreatic cancer development [85]. In chronic inflammatory conditions such as obesity-related pancreatitis or diabetes, sustained NF-κB activation disrupts the PI3K/Akt/mTOR signaling pathway. This disruption can promote aberrant cell growth, survival, and metabolic alterations in pancreatic cells, contributing significantly to the initiation and progression of pancreatic cancer. Thus, understanding and targeting these inflammatory-mediated mechanisms are critical for developing strategies to mitigate pancreatic disease progression associated with PI3K/Akt/mTOR pathway dysregulation [86].

##### Resultant dysregulation in pancreatic cell fate predisposing pancreas to pancreatic adenocarcinoma

The NF-κB pathway’s influence on the PI3K/Akt/mTOR signaling pathway disrupts pancreatic cell fate and predisposes the pancreas to pancreatic adenocarcinoma through interconnected mechanisms [87]. Chronic inflammation leads to sustained NF-κB activation, which profoundly impacts PI3K/Akt/mTOR signaling by altering the expression and activity of pathway components. This dysregulation impairs the differentiation of pancreatic progenitor cells, potentially resulting in the formation of abnormal cell types or progenitors that fail to mature into functional pancreatic cells, both endocrine and exocrine [88]. Moreover, NF-κB activation enhances PI3K isoforms such as PI3Kα and PI3Kβ, boosting phosphatidylinositol 3,4,5-trisphosphate (PIP3) production and subsequent Akt activation. Activated Akt, in turn, drives cell proliferation by phosphorylating targets involved in cell cycle progression while suppressing apoptotic pathways [89]. This survival advantage extends to genetically altered or damaged cells, promoting their persistence and increasing susceptibility to malignant transformation under chronic inflammatory conditions. Additionally, dysregulated PI3K/Akt/mTOR signaling induces metabolic reprogramming in pancreatic cells, favoring aerobic glycolysis (the Warburg effect) to meet the energy demands of rapidly proliferating cancer cells. This metabolic shift not only supports cell growth but also enhances cell survival under stressful conditions, contributing to tumor progression [90]. In terms of predisposing the pancreas to pancreatic adenocarcinoma, NF-κB-mediated dysregulation of PI3K/Akt/mTOR signaling induces epithelial-mesenchymal transition (EMT) in pancreatic epithelial cells. EMT empowers cancer cells with migratory and invasive properties, facilitating metastasis to distant organs. Furthermore, dysregulated PI3K/Akt/mTOR signaling promotes uncontrolled cell growth and survival, fostering the development of pre-cancerous lesions like pancreatic intraepithelial neoplasia (PanIN), which can progress into invasive pancreatic adenocarcinoma. Within the tumor microenvironment, dysregulated PI3K/Akt/mTOR signaling may suppress the anti-tumor immune response, allowing cancer cells to evade detection and destruction by immune cells. This immune evasion, coupled with the pro-inflammatory milieu fostered by NF-κB activation and dysregulated PI3K/Akt/mTOR signaling, supports tumor growth, angiogenesis, and metastasis [91].

#### 8. JAK/STAT Signaling Pathway

##### NF-κB in disrupting this signaling pathway in pancreas

The JAK/STAT signaling pathway is crucial in pancreas development, governing critical processes like cell proliferation, differentiation, and immune responses [92]. Dysregulation of this pathway, driven by inflammation and mediated notably by NF-κB, disrupts pancreatic homeostasis and contributes to various pancreatic diseases. Several mechanisms illustrate how NF-κB-mediated inflammation can disrupt the gene expression of the JAK/STAT signaling pathway [93]. Inflammatory cytokines such as IL-6, IL-1β, and TNF-α activate NF-κB, directly impacting JAK/STAT pathway components. NF-κB binds to κB sites in the promoters of genes encoding JAK family members (e.g., JAK1, JAK2, JAK3), STAT transcription factors (e.g., STAT1, STAT3, STAT5), and downstream targets like SOCS and CIS, thereby modulating their transcriptional activity [94]. This regulation can lead to both upregulation and downregulation of these components, altering pathway dynamics crucial for pancreatic cell fate and immune responses. Furthermore, inflammation induces oxidative stress, influencing the stability and function of JAK/STAT pathway components. Reactive oxygen species (ROS) affect the phosphorylation status of JAKs and STATs, thereby modifying their signaling activities. Epigenetic modifications induced by inflammation, such as changes in DNA methylation and histone modifications, also play a role [95]. These alterations impact the accessibility of JAK/STAT pathway gene promoters, potentially leading to aberrant expression of JAKs, STATs, and their regulated genes under inflammatory conditions. Inflammatory signals additionally stimulate specific miRNAs that target mRNAs encoding JAK/STAT pathway components, promoting their degradation or translational repression. This miRNA-mediated regulation can directly inhibit JAKs, STATs, or negative regulators like SOCS proteins, thereby modulating pathway activity and influencing pancreatic cell responses to inflammation and disease progression. NF-κB activation not only interacts directly with JAK/STAT pathway genes but also cross-talks with other signaling pathways such as MAPK and PI3K/Akt. This interplay modulates the activity and expression of JAK/STAT pathway components, influencing pancreatic cell fate decisions, immune responses, and the development of pancreatic diseases. For example, in conditions like chronic pancreatitis or pancreatic ductal adenocarcinoma (PDAC), sustained NF-κB activation dysregulates JAK/STAT signaling [96]. Dysregulated STAT3 signaling, in particular, can promote cell proliferation, survival, and inflammation within pancreatic cells, thereby contributing significantly to tumor growth and disease progression. This dysregulation underscores the critical role of NF-κB-mediated inflammation in driving pathological changes in the JAK/STAT pathway that ultimately impact pancreatic health and disease outcomes [97].

##### Resultant dysregulation in pancreatic cell fate predisposing pancreas to pancreatic adenocarcinoma

NF-κB exerts significant influence on the JAK/STAT signaling pathway, disrupting pancreatic cell fate and predisposing the pancreas to pancreatic adenocarcinoma through interconnected mechanisms [98]. Chronic inflammation triggers sustained NF-κB activation, which dysregulates the JAK/STAT pathway by altering the expression and activity of JAK kinases and STAT transcription factors. This dysregulation impairs the differentiation of pancreatic progenitor cells, potentially leading to the formation of abnormal cell types or inadequate differentiation into mature pancreatic cells, both endocrine and exocrine [99]. Moreover, NF-κB activation enhances the production of pro-inflammatory cytokines like IL-6 and IL-1β, which in turn activate JAK/STAT signaling pathways, particularly STAT3. Increased STAT activation promotes a chronic inflammatory microenvironment within the pancreas, fostering conditions favorable for tumorigenesis [100]. This inflammatory milieu not only supports tumor growth but also facilitates angiogenesis and metastasis through its pro-inflammatory effects. In the context of pancreatic adenocarcinoma, dysregulated JAK/STAT signaling under NF-κB influence plays a key role in enhancing tumorigenesis and disease progression. Activated STAT proteins, especially STAT3, drive pathways involved in cell proliferation and survival. They induce the expression of genes critical for cell cycle progression and anti-apoptotic mechanisms, thereby promoting tumor cell survival and uncontrolled growth [101]. Additionally, dysregulated JAK/STAT signaling contributes to the acquisition of metastatic potential in pancreatic cancer cells by inducing epithelial-mesenchymal transition (EMT), a process that enhances their invasive and metastatic capabilities. Furthermore, this dysregulation confers resistance to therapeutic interventions such as chemotherapy and targeted therapies. STAT3 activation, for instance, upregulates anti-apoptotic proteins and drug efflux pumps, thereby reducing the efficacy of treatments aimed at inhibiting tumor growth and progression. In chronic inflammatory conditions like pancreatitis or obesity-related inflammation, sustained NF-κB activation disrupts the JAK/STAT signaling pathway, particularly enhancing STAT3 activation. This perpetuates a pro-inflammatory and pro-tumorigenic microenvironment in the pancreas, where persistent STAT3 activation drives mechanisms of cell proliferation, survival, and immune evasion [102]. These processes collectively contribute to the initiation, progression, and therapeutic resistance observed in pancreatic adenocarcinoma, highlighting the critical interplay between NF-κB-mediated inflammation and dysregulated JAK/STAT signaling in pancreatic cancer development [103, 104, 105].

#### 9. Hippo Signaling Pathway

##### NF-κB in disrupting this signaling pathway in pancreas

The Hippo signaling pathway, crucial for pancreas development, regulates organ size, cell proliferation, and tissue homeostasis [106]. Inflammation, particularly mediated by NF-κB, can disrupt this pathway, influencing pancreatic development and contributing to disease states. NF-κB-mediated inflammation can dysregulate the Hippo pathway through several mechanisms [107]. NF-κB activation directly influences the expression of Hippo pathway components such as YAP (Yes-associated protein) and TAZ (transcriptional coactivator with PDZ-binding motif). This activation can upregulate YAP/TAZ or alter the expression of upstream regulators like MST1/2 and LATS1/2 by binding to their promoters [108]. Additionally, NF-κB induces cytokine production (e.g., TNF-α, IL-1β), which can modulate YAP/TAZ nuclear localization and activity, thereby affecting downstream gene expression in pancreatic cells. Oxidative stress induced by NF-κB-mediated inflammation generates reactive oxygen species (ROS) that impact Hippo pathway kinases (e.g., MST1/2, LATS1/2). ROS-mediated oxidative stress alters the phosphorylation and regulation of YAP/TAZ, potentially promoting their nuclear localization independent of MST/LATS kinase activity [109]. Furthermore, inflammation-driven changes in DNA methylation patterns and histone modifications can modify accessibility to Hippo pathway gene promoters, thereby influencing pathway activity in pancreatic cells. Inflammatory signals also induce specific microRNAs (miRNAs) targeting mRNAs encoding Hippo pathway components. Dysregulated miRNA expression profiles under NF-κB influence may post-transcriptionally repress Hippo pathway regulators or effectors, thereby modulating pathway activity and cellular responses [110]. NF-κB activation enhances YAP/TAZ activity by promoting their nuclear translocation and transcriptional activity. This influence can alter phosphorylation status or induce cytokines that affect their regulation, subsequently activating pro-survival pathways downstream of YAP/TAZ. Moreover, NF-κB interacts with other signaling pathways (e.g., Wnt, Notch) that converge on the Hippo pathway, potentially amplifying YAP/TAZ signaling or altering cellular sensitivity to Hippo pathway regulation under inflammatory conditions. In chronic pancreatitis or pancreatic cancer, sustained NF-κB activation dysregulates the Hippo signaling pathway. Enhanced YAP/TAZ activity promotes cell proliferation, inhibits apoptosis, and induces epithelial-mesenchymal transition (EMT) in pancreatic epithelial cells, all contributing to tumor initiation and progression. This dysregulation alters pancreatic cell fate and creates a microenvironment favorable for tumor growth [111].

##### Resultant dysregulation in pancreatic cell fate predisposing pancreas to pancreatic adenocarcinoma

The impact of NF-κB on the Hippo signaling pathway can disrupt pancreatic cell fate and predispose the pancreas to pancreatic adenocarcinoma through several interconnected mechanisms. NF-κB-mediated inflammation can lead to the activation and nuclear translocation of YAP (Yes-associated protein) and TAZ (transcriptional coactivator with PDZ-binding motif), key effectors of the Hippo pathway [112]. This activation promotes cell proliferation by enhancing the expression of genes involved in cell cycle progression and inhibiting genes that regulate cell differentiation. Increased YAP/TAZ activity under NF-κB influence may suppress the expression of differentiation markers in pancreatic progenitor cells, leading to the accumulation of undifferentiated cells or cells with altered differentiation patterns, which are more prone to malignant transformation [113]. Additionally, NF-κB activation and subsequent YAP/TAZ activation can inhibit apoptosis in pancreatic cells by upregulating anti-apoptotic genes. This anti-apoptotic effect allows damaged or genetically altered cells to survive, contributing to tumor initiation and progression. Dysregulated Hippo signaling, characterized by increased YAP/TAZ activity under NF-κB influence, promotes cell proliferation, survival, and epithelial-mesenchymal transition (EMT) in pancreatic cells. EMT endows cancer cells with invasive properties and facilitates metastasis to distant organs [114]. Moreover, YAP/TAZ activation in pancreatic adenocarcinoma cells can confer resistance to chemotherapy and targeted therapies, reducing treatment efficacy and contributing to disease progression. NF-κB activation induces the production of pro-inflammatory cytokines and chemokines that create a favorable microenvironment for tumor growth. This inflammatory milieu supports angiogenesis, immune evasion, and enhances the survival of cancer cells in the pancreatic tumor microenvironment. In chronic inflammation associated with conditions such as pancreatitis or obesity-related inflammation, sustained NF-κB activation dysregulates the Hippo signaling pathway [115]. Enhanced YAP/TAZ activity promotes uncontrolled cell proliferation, inhibits apoptosis, and facilitates EMT in pancreatic epithelial cells, leading to the initiation and progression of pancreatic adenocarcinoma [116].

## Discussion

### Landscape of Dysregulations in Pancreatic Signaling Pathways Induced by NF-κB

NF-κB-mediated inflammation profoundly impacts pancreatic signaling pathways, tipping the balance towards a pathologic state characterized by dysregulated cellular processes and increased susceptibility to pancreatic adenocarcinoma. NF-κB activation disrupts the Notch signaling pathway, impairing cell differentiation and promoting survival mechanisms that drive oncogenesis. It induces aberrant Wnt signaling, leading to uncontrolled cell proliferation and inhibition of differentiation, which are crucial for tumor initiation. NF-κB inhibits the Hippo signaling pathway, resulting in the hyperactivation of YAP/TAZ, which enhances cell proliferation, inhibits apoptosis, and facilitates epithelial-mesenchymal transition (EMT), promoting tumor progression. NF-κB-driven cytokine production activates the JAK/STAT signaling pathway, fostering a pro-inflammatory environment that supports tumor growth and immune evasion. NF-κB alters TGF-β signaling, impacting pancreatic cell differentiation and promoting fibrosis, contributing to tumor microenvironment remodeling. It disrupts the BMP signaling pathway, affecting pancreatic development and beta-cell function, potentially leading to metabolic dysregulation and cancer progression. NF-κB influences FGF signaling, affecting pancreatic progenitor cell proliferation and differentiation, which are crucial for pancreatic tissue homeostasis. NF-κB activation promotes the PI3K/Akt/mTOR pathway activity, enhancing cell survival and metabolism, thereby contributing to tumor cell proliferation and resistance to therapy.

### Consequences

#### Cell Fate and Differentiation

NF-κB-induced dysregulation of signaling pathways impairs pancreatic cell differentiation, promoting the accumulation of undifferentiated cells susceptible to oncogenic transformation.

#### Tumor Microenvironment

Chronic inflammation mediated by NF-κB creates a pro-tumorigenic microenvironment characterized by enhanced cell survival, angiogenesis, and immune evasion.

#### Metastasis and Therapy Resistance

Dysregulated pathways under NF-κB influence promote epithelial-mesenchymal transition (EMT), metastatic spread, and resistance to conventional therapies, complicating treatment outcomes.

NF-κB-induced damage disrupts the delicate balance between pancreatic proliferation and differentiation by dysregulating key signaling pathways critical for these processes.

### Impact on Proliferation

#### 1. Wnt Signaling Pathway

##### NF-κB Influence

Activates Wnt signaling, promoting cell proliferation.

##### Effect

Enhances beta-cell proliferation but inhibits differentiation, contributing to pancreatic hyperplasia and potential tumorigenesis.

#### 2. PI3K/Akt/mTOR Signaling Pathway

##### NF-κB Influence

Activates PI3K/Akt/mTOR pathway, stimulating cell growth and proliferation.

##### Effect

Promotes beta-cell survival and proliferation but may suppress differentiation, exacerbating cell fate imbalance under inflammatory conditions.

#### 3. JAK/STAT Signaling Pathway

##### NF-κB Influence

Induces pro-inflammatory cytokines that activate JAK/STAT signaling.

##### Effect

Enhances beta-cell proliferation and immune modulation but may suppress differentiation pathways, contributing to dysregulated pancreatic function.

### Impact on Differentiation

#### 1. Notch Signaling Pathway

##### NF-κB Influence

Disrupts Notch signaling, crucial for cell fate determination and differentiation.

##### Effect

Impairs endocrine and exocrine cell differentiation, leading to the accumulation of undifferentiated cells prone to oncogenic transformation.

#### 2. Hippo Signaling Pathway

##### NF-κB Influence

Inhibits Hippo pathway, activating YAP/TAZ transcription co-activators.

##### Effect

Suppresses pancreatic cell differentiation and enhances cell proliferation, promoting tumorigenesis and impairing tissue homeostasis.

#### 3. TGF-β Signaling Pathway

##### NF-κB Influence

Modulates TGF-β signaling, affecting cell differentiation and fibrosis.

##### Effect

Alters pancreatic progenitor cell differentiation and may contribute to fibrotic changes that disrupt tissue architecture and function.

NF-κB-induced damage skews the balance towards proliferation over differentiation in pancreatic cells through several mechanisms:

### Enhanced Proliferation

Activation of pathways like Wnt and PI3K/Akt/mTOR promotes cell growth and survival, crucial for tissue repair but may lead to hyperplasia or oncogenic transformation when dysregulated.

### Impaired Differentiation

Disruption of Notch, Hippo, and TGF-β pathways inhibits proper cell differentiation, leading to the accumulation of immature or undifferentiated cells that are susceptible to tumorigenesis.

### Clinical Implications

The findings of research into NF-κB-induced damage on pancreatic signaling pathways have significant clinical implications for the management and treatment of pancreatic diseases, particularly pancreatic adenocarcinoma. Developing targeted therapies that inhibit NF-κB activation or downstream signaling pathways implicated in pancreatic cancer pathogenesis could potentially disrupt the pro-tumorigenic effects of NF-κB and restore normal signaling pathway activity. Exploring combination therapies that target both NF-κB and specific dysregulated signaling pathways, such as Wnt and PI3K/Akt/mTOR, may enhance treatment efficacy and overcome resistance mechanisms observed in pancreatic cancer. Identifying biomarkers associated with NF-κB activation and dysregulated signaling pathways could improve early detection, prognostication, and personalized treatment strategies for patients with pancreatic adenocarcinoma. Investigating immunotherapy approaches that target NF-κB-mediated inflammation to modulate the tumor microenvironment and enhance anti-tumor immune responses may potentially improve outcomes for patients with advanced pancreatic cancer. Implementing precision medicine approaches that consider the molecular and genetic profiles of pancreatic tumors, including NF-κB pathway activation status, could help tailor treatment strategies and improve therapeutic outcomes based on individual patient characteristics. Conducting clinical trials focused on new therapeutic strategies targeting NF-κB and dysregulated signaling pathways identified in preclinical research could translate these findings into effective clinical interventions for pancreatic cancer patients. Integrating insights from NF-κB-induced damage research into clinical practice guidelines for the management and follow-up of patients at high risk for pancreatic adenocarcinoma can ensure timely intervention and personalized care plans. Translating the research findings into clinical implications involves leveraging new insights into NF-κB-mediated signaling dysregulation to develop therapeutic approaches, improve patient outcomes, and advance the field of pancreatic cancer treatment.

## Conclusions

This study looks into how NF-κB-mediated inflammation damages signaling pathways critical for pancreas homeostasis, thereby increasing the risk of pancreatic adenocarcinoma. NF-κB activation plays a key role in orchestrating a cascade of molecular events that disrupt key signaling pathways involved in pancreatic development, cell fate determination, and tumor suppression mechanisms. NF-κB exerts its deleterious effects by dysregulating several crucial signaling pathways in the pancreas. NF-κB activation can perturb the Notch signaling pathway, disrupting cell differentiation and promoting cell survival pathways that contribute to tumor initiation. Dysregulated Wnt signaling under NF-κB influence promotes aberrant cell proliferation and blocks differentiation, fostering a tumorigenic environment in the pancreas. NF-κB-mediated activation of YAP/TAZ via Hippo pathway inhibition promotes uncontrolled cell proliferation, inhibits apoptosis, and enhances epithelial-mesenchymal transition, facilitating tumor progression. Chronic NF-κB activation induces cytokine production that stimulates the JAK/STAT signaling pathway, promoting inflammation and creating a microenvironment conducive to tumor growth. Furthermore, NF-κB dysregulation impacts other signaling pathways, such as TGF-β, BMP, FGF, and PI3K/Akt/mTOR, influencing pancreatic cell fate, differentiation, and survival, thereby predisposing the pancreas to oncogenic transformation. The cumulative effect of NF-κB-mediated damage to these signaling pathways increases the susceptibility of pancreatic cells to oncogenic transformation and the development of pancreatic adenocarcinoma. Persistent inflammation, as seen in chronic pancreatitis or obesity-related conditions, sustains NF-κB activation, perpetuating the dysregulation of these pathways and promoting tumorigenesis. Targeting NF-κB and its downstream signaling pathways represents a promising therapeutic strategy to mitigate inflammation-driven pancreatic damage and improve clinical outcomes for patients with pancreatic cancer.

## Abbreviations

NF-κB: Nuclear Factor kappa-light-chain-enhancer of activated B cells
Wnt: Wingless/Integrated
SHH: Sonic Hedgehog
FGF: Fibroblast Growth Factor
TGF-β: Transforming Growth Factor-beta
BMP: Bone Morphogenetic Protein
PI3K: Phosphoinositide 3-Kinase
Akt: Protein Kinase B
mTOR: Mammalian Target of Rapamycin
JAK: Janus Kinase
STAT: Signal Transducer and Activator of Transcription
YAP: Yes-associated Protein
TAZ: Transcriptional co-Activator with PDZ-binding motif
EMT: Epithelial-Mesenchymal Transition

## Declarations

### Ethics declarations

**Ethics approval and consent to participate**

Not applicable.

### Consent for publication

Not applicable.

### Data Availability statement

All data generated or analyzed during this study are included in this article.

## Acknowledgements

“Not applicable”.

## Authors’ Information

1. **Ovais Shafi (OS)\*** is the author of the study and was involved in the idea, concept, design, and methodology of the study, literature search and references. He did the writing, editing, and revision of the manuscript. He was involved in drawing the findings, results, conclusions, implications of the study, interpretation of the data and was involved in all aspects of the study. He prepared and wrote discussion, results, conclusions and all areas of the study. OS extracted and analyzed the data. He was involved in critical evaluation, audit of every aspect of the study, data extraction, adherence of the study to relevant PRISMA guidelines, limitations of the study, references, and all others. He was involved in drawing PRISMA Flow Diagram. The author read and approved the manuscript. He investigated the pancreatic signaling pathways for their roles in relation to this study: Notch signaling pathway, Wnt signaling pathway, Shh signaling pathway, FGF signaling pathway, TGF-β signaling pathway, BMP signaling pathway, PI3K/Akt/mTOR signaling pathway, JAK/STAT signaling pathway, Hippo signaling pathway. **Ovais Shafi (OS)\***, MBBS - Sindh Medical College - Dow University of Health Sciences, Karachi, Pakistan. He aspires to become an eminent ‘Physician Scientist’. He is devoted to the research in disease development mechanisms, disease origins and therapeutics. OS is also passionate about multiple research areas including clinical trials, clinical medicine, therapeutics, regenerative medicine, precision medicine including gene therapies, finding disease specific targets for gene therapy, role of disease genomics and epigenetics in diagnosis, management, and therapeutics development. He is dedicated to the field of research and clinical medicine. Email address*: **Corresponding author: OS** Correspondence to <u>Ovais Shafi</u>
2. **Rahimeen Rajpar (RR)** is also the author of the study and contributed to the writing, editing and revision of the study. She also contributed to the results and conclusions of this manuscript along with working on the findings. She contributed to investigating the pancreatic signaling pathways for their roles in relation to this study: Notch signaling pathway, Wnt signaling pathway, Shh signaling pathway, FGF signaling pathway, TGF-β signaling pathway, BMP signaling pathway, PI3K/Akt/mTOR signaling pathway, JAK/STAT signaling pathway, Hippo signaling pathway. Rahimeen Rajpar, MD is a dedicated medical professional with a passion for unraveling the mysteries of disease origins and progression. Currently pursuing her residency in internal medicine. RR is committed to advancing the field of medical research. Her goal is to become a leader in the field of Medicine, looking at the medical intricacies from a different lens. Apart from research RR remains dedicated to improving the lives of her patients through comprehensive care and by working towards scientific discoveries. RR is a MBBS graduate from Sindh Medical College – Jinnah Sindh Medical University, Karachi, Pakistan.
3. **Aakash (AA)** is the co-author of the study. He contributed to the results and conclusions of the study, also contributed to the writing and editing of these sections. He also contributed to the references. He contributed to investigating the pancreatic signaling pathways for their roles in relation to this study: Notch signaling pathway, Wnt signaling pathway, Shh signaling pathway, FGF signaling pathway, TGF-β signaling pathway, BMP signaling pathway, PI3K/Akt/mTOR signaling pathway, JAK/STAT signaling pathway, Hippo signaling pathway. Aakash MD, currently working as PGY1 in Florida State University Cape Coral Hospital and he is MBBS from Sindh Medical College - Dow University of Health Sciences, Karachi, Pakistan. He is passionate about pursuing fellowship in gastroenterology. His goal is also to translate emerging findings in research of disease development mechanisms/ origins into their clinical implications.
4. **Muhammad Waqas (MW)** is the co-author of the study. He contributed to the results and conclusions of the study, also contributed to the writing and editing of these sections. He also contributed to the references. He contributed to investigating the pancreatic signaling pathways for their roles in relation to this study: Notch signaling pathway, Wnt signaling pathway, Shh signaling pathway, FGF signaling pathway, TGF-β signaling pathway, BMP signaling pathway, PI3K/Akt/mTOR signaling pathway, JAK/STAT signaling pathway, Hippo signaling pathway. Muhammad Waqas, MBBS - Sindh Medical College - Dow University of Health Sciences, Karachi, Pakistan. He is ECFMG Certified. His future goals include residency in Internal Medicine/Neurology, and fellowship in Cardiology/Nephrology/Critical Care.
5. **Madiha Haseeb (MH)** is the co-author of the study. She contributed to the results of the study, also contributed to its writing and editing. She contributed to investigating the pancreatic signaling pathways for their roles in relation to this study: Notch signaling pathway, Wnt signaling pathway, Shh signaling pathway, FGF signaling pathway, TGF-β signaling pathway, BMP signaling pathway, PI3K/Akt/mTOR signaling pathway, JAK/STAT signaling pathway, Hippo signaling pathway. Madiha Haseeb, MBBS - Sindh Medical College - Dow University of Health Sciences, Karachi, Pakistan. She has completed her all USMLE examinations and is ECFMG certified. Her areas of interest include internal medicine and neurology. Her future goal includes being a certified internal medicine specialist.
6. **Raveena (RA)** is the co-author of the study. She contributed to the results and conclusions of the study, also contributed to the writing and editing of these sections. She contributed to investigating the pancreatic signaling pathways for their roles in relation to this study: Notch signaling pathway, Wnt signaling pathway, Shh signaling pathway, FGF signaling pathway, TGF-β signaling pathway, BMP signaling pathway, PI3K/Akt/mTOR signaling pathway, JAK/STAT signaling pathway, Hippo signaling pathway. Raveena, MBBS - Sindh Medical College – Jinnah Sindh Medical University, Karachi, Pakistan. She is passionate about research in surgery and disease development mechanisms including neurodegenerative diseases, oncogenesis and others. She is ECFMG Certified. She is passionate about residency in Internal Medicine/Surgery. Her goal is to make significant impact in the field of Research.
7. **Madhurta Kumari (MAK)** is the co-author of the study. She contributed to the results of the study, also contributed to its writing and editing. She contributed to investigating the pancreatic signaling pathways for their roles in relation to this study: Notch signaling pathway, Wnt signaling pathway, Shh signaling pathway, FGF signaling pathway, TGF-β signaling pathway, BMP signaling pathway, PI3K/Akt/mTOR signaling pathway, JAK/STAT signaling pathway, Hippo signaling pathway. Madhurta Kumari, MBBS - Chandka Medical College – SMBBMU, Larkana Sindh, Pakistan. She is ECFMG certified and her future goals include fellowship in Gastroenterology.
8. **Ajay Kumar (AK)** is the co-author of the study. He contributed to the results of the study, also contributed to its writing and editing. He contributed to investigating the pancreatic signaling pathways for their roles in relation to this study: Notch signaling pathway, Wnt signaling pathway, Shh signaling pathway, FGF signaling pathway, TGF-β signaling pathway, BMP signaling pathway, PI3K/Akt/mTOR signaling pathway, JAK/STAT signaling pathway, Hippo signaling pathway. Ajay Kumar, MBBS - Chandka Medical College – SMBBMU, Larkana Sindh, Pakistan. He is ECFMG certified and his future goals include fellowship in Cardiology.
9. **Muskan Kumari (MUK)** is the co-author of the study. She contributed to the results of the study, also contributed to its writing and editing. She contributed to investigating the pancreatic signaling pathways for their roles in relation to this study: Notch signaling pathway, Wnt signaling pathway, Shh signaling pathway, FGF signaling pathway, TGF-β signaling pathway, BMP signaling pathway, PI3K/Akt/mTOR signaling pathway, JAK/STAT signaling pathway, Hippo signaling pathway. Muskan Kumari, MBBS - Chandka Medical College – SMBBMU, Larkana Sindh, Pakistan. She is ECFMG certified and her future goals include fellowship in Cardiology.
10. **Muhammad Danial Yaqub (MDY)** is the co-author of the study. He contributed to the results of the study. He contributed to investigating the pancreatic signaling pathways for their roles in relation to this study: Notch signaling pathway, Wnt signaling pathway, Shh signaling pathway, FGF signaling pathway, TGF-β signaling pathway, BMP signaling pathway, PI3K/Akt/mTOR signaling pathway, JAK/STAT signaling pathway, Hippo signaling pathway. Muhammad Danial Yaqub, MBBS - Dow University of Health Sciences, Karachi, Pakistan. He is currently working as a Trust Grade Doctor at Lincoln County Hospital, Lincoln, United Kingdom. He has keen interests in Gastroenterology and Clinical Medicine. MDY aspires to be a researcher which can present new data which can pave the way for early diagnosis and treatment of gastrointestinal diseases. His goal is also to translate emerging findings in research of disease development mechanisms/ origins into their clinical implications.

*The work and contributions of everyone have been described in detail, the order is randomized and the numbering is just for referencing purpose*.

## References

1. Huang H, Liu Y, Daniluk J, Gaiser S, Chu J, Wang H, Li ZS, Logsdon CD, Ji B. Activation of nuclear factor-κB in acinar cells increases the severity of pancreatitis in mice. Gastroenterology. 2013 Jan;144(1):202–10. doi: 10.1053/j.gastro.2012.09.059. Epub 2012 Oct 3. PMID: 23041324; PMCID: PMC3769090.

2. Jakkampudi A, Jangala R, Reddy BR, Mitnala S, Nageshwar Reddy D, Talukdar R. NF-κB in acute pancreatitis: Mechanisms and therapeutic potential. Pancreatology. 2016 Jul-Aug;16(4):477–88. doi: 10.1016/j.pan.2016.05.001. Epub 2016 May 24. PMID: 27282980.

3. Wu N, Xu XF, Xin JQ, Fan JW, Wei YY, Peng QX, Duan LF, Wang W, Zhang H. The effects of nuclear factor-kappa B in pancreatic stellate cells on inflammation and fibrosis of chronic pancreatitis. J Cell Mol Med. 2021 Feb;25(4):2213–2227. doi: 10.1111/jcmm.16213. Epub 2020 Dec 30. PMID: 33377616; PMCID: PMC7882951.

4. Meyerovich K, Fukaya M, Terra LF, Ortis F, Eizirik DL, Cardozo AK. The non-canonical NF-κB pathway is induced by cytokines in pancreatic beta cells and contributes to cell death and proinflammatory responses in vitro. Diabetologia. 2016 Mar;59(3):512–21. doi: 10.1007/s00125-015-3817-z. Epub 2015 Dec 3. PMID: 26634571.

5. Zhuo F, Luo S, He W, Feng Z, Hu Y, Xu J, Wang Z, Xu J. The Role of Signaling Pathways in Pancreatic Cancer Targeted Therapy. Am J Clin Oncol. 2023 Mar 1;46(3):121–128. doi: 10.1097/COC.0000000000000979. Epub 2023 Feb 3. PMID: 36735511.

6. Jones S, Zhang X, Parsons DW, Lin JC, Leary RJ, Angenendt P, Mankoo P, Carter H, Kamiyama H, Jimeno A, Hong SM, Fu B, Lin MT, Calhoun ES, Kamiyama M, Walter K, Nikolskaya T, Nikolsky Y, Hartigan J, Smith DR, Hidalgo M, Leach SD, Klein AP, Jaffee EM, Goggins M, Maitra A, Iacobuzio-Donahue C, Eshleman JR, Kern SE, Hruban RH, Karchin R, Papadopoulos N, Parmigiani G, Vogelstein B, Velculescu VE, Kinzler KW. Core signaling pathways in human pancreatic cancers revealed by global genomic analyses. Science. 2008 Sep 26;321(5897):1801-6. doi: 10.1126/science.1164368. Epub 2008 Sep 4. PMID: 18772397; PMCID: PMC2848990.

7. Song G, Zhang Y, Jiang Y, Zhang H, Gu W, Xu X, Yao J, Chen Z. Signaling pathway of targeting the pancreas in the treatment of diabetes under the precision medicine big data evaluation system. Front Genet. 2023 Feb 17;14:1119181. doi: 10.3389/fgene.2023.1119181. PMID: 36873938; PMCID: PMC9981801.

8. Reddy SA. Signaling pathways in pancreatic cancer. Cancer J. 2001 Jul-Aug;7(4):274–86. PMID: 11561604.

9. Maniati E, Bossard M, Cook N, Candido JB, Emami-Shahri N, Nedospasov SA, Balkwill FR, Tuveson DA, Hagemann T. Crosstalk between the canonical NF-κB and Notch signaling pathways inhibits Pparγ expression and promotes pancreatic cancer progression in mice. J Clin Invest. 2011 Dec;121(12):4685–99. doi: 10.1172/JCI45797. Epub 2011 Nov 7. PMID: 22056382; PMCID: PMC3225987.

10. Pramanik KC, Makena MR, Bhowmick K, Pandey MK. Advancement of NF-κB Signaling Pathway: A Novel Target in Pancreatic Cancer. Int J Mol Sci. 2018 Dec 5;19(12):3890. doi: 10.3390/ijms19123890. PMID: 30563089; PMCID: PMC6320793.

11. Prabhu L, Mundade R, Korc M, Loehrer PJ, Lu T. Critical role of NF-κB in pancreatic cancer. Oncotarget. 2014 Nov 30;5(22):10969–75. doi: 10.18632/oncotarget.2624. PMID: 25473891; PMCID: PMC4294354.

12. Osipo C, Golde TE, Osborne BA, Miele LA. Off the beaten pathway: the complex cross talk between Notch and NF-kappaB. Lab Invest. 2008 Jan;88(1):11–7. doi: 10.1038/labinvest.3700700. Epub 2007 Dec 3. PMID: 18059366.

13. Thomas MM, Zhang Y, Mathew E, Kane KT, Maillard I, Pasca di Magliano M. Epithelial Notch signaling is a limiting step for pancreatic carcinogenesis. BMC Cancer. 2014 Nov 22;14:862. doi: 10.1186/1471-2407-14-862. PMID: 25416148; PMCID: PMC4289235.

14. Siveke JT, Lubeseder-Martellato C, Lee M, Mazur PK, Nakhai H, Radtke F, Schmid RM. Notch signaling is required for exocrine regeneration after acute pancreatitis. Gastroenterology. 2008 Feb;134(2):544–55. doi: 10.1053/j.gastro.2007.11.003. Epub 2007 Nov 4. PMID: 18242220.

15. Chung WC, Xu K. Notch signaling pathway in pancreatic tumorigenesis. Adv Cancer Res. 2023;159:1–36. doi: 10.1016/bs.acr.2023.02.001. Epub 2023 Feb 28. PMID: 37268393.

16. Li XY, Zhai WJ, Teng CB. Notch Signaling in Pancreatic Development. Int J Mol Sci. 2015 Dec 30;17(1):48. doi: 10.3390/ijms17010048. PMID: 26729103; PMCID: PMC4730293.

17. Avila JL, Kissil JL. Notch signaling in pancreatic cancer: oncogene or tumor suppressor? Trends Mol Med. 2013 May;19(5):320–7. doi: 10.1016/j.molmed.2013.03.003. Epub 2013 Mar 29. PMID: 23545339; PMCID: PMC3648591.

18. Mysliwiec P, Boucher MJ. Targeting Notch signaling in pancreatic cancer patients--rationale for new therapy. Adv Med Sci. 2009;54(2):136–42. doi: 10.2478/v10039-009-0026-3. PMID: 19758972.

19. Yan W, Menjivar RE, Bonilla ME, Steele NG, Kemp SB, Du W, Donahue KL, Brown KL, Carpenter ES, Avritt FR, Irizarry-Negron VM, Yang S, Burns WR 3rd, Zhang Y, Pasca di Magliano M, Bednar F. Notch Signaling Regulates Immunosuppressive Tumor-Associated Macrophage Function in Pancreatic Cancer. Cancer Immunol Res. 2024 Jan 3;12(1):91–106. doi: 10.1158/2326-6066.CIR-23-0037. PMID: 37931247; PMCID: PMC10842043.

20. Napolitano T, Silvano S, Ayachi C, Plaisant M, Sousa-Da-Veiga A, Fofo H, Charles B, Collombat P. Wnt Pathway in Pancreatic Development and Pathophysiology. Cells. 2023 Feb 9;12(4):565. doi: 10.3390/cells12040565. PMID: 36831232; PMCID: PMC9954665.

21. Napolitano T, Silvano S, Ayachi C, Plaisant M, Sousa-Da-Veiga A, Fofo H, Charles B, Collombat P. Wnt Pathway in Pancreatic Development and Pathophysiology. Cells. 2023 Feb 9;12(4):565. doi: 10.3390/cells12040565. PMID: 36831232; PMCID: PMC9954665.

22. Sharon N, Vanderhooft J, Straubhaar J, Mueller J, Chawla R, Zhou Q, Engquist EN, Trapnell C, Gifford DK, Melton DA. Wnt Signaling Separates the Progenitor and Endocrine Compartments during Pancreas Development. Cell Rep. 2019 May 21;27(8):2281–2291.e5. doi: 10.1016/j.celrep.2019.04.083. PMID: 31116975; PMCID: PMC6933053.

23. Sano M, Driscoll DR, DeJesus-Monge WE, Quattrochi B, Appleman VA, Ou J, Zhu LJ, Yoshida N, Yamazaki S, Takayama T, Sugitani M, Nemoto N, Klimstra DS, Lewis BC. Activation of WNT/β-Catenin Signaling Enhances Pancreatic Cancer Development and the Malignant Potential Via Up-regulation of Cyr61. Neoplasia. 2016 Dec;18(12):785–794. doi: 10.1016/j.neo.2016.11.004. Epub 2016 Nov 25. PMID: 27889647; PMCID: PMC5126137.

24. Hawkins HJ, Yacob BW, Brown ME, Goldstein BR, Arcaroli JJ, Bagby SM, Hartman SJ, Macbeth M, Goodspeed A, Danhorn T, Lentz RW, Lieu CH, Leal AD, Messersmith WA, Dempsey PJ, Pitts TM. Examination of Wnt signaling as a therapeutic target for pancreatic ductal adenocarcinoma (PDAC) using a pancreatic tumor organoid library (PTOL). PLoS One. 2024 Apr 10;19(4):e0298808. doi: 10.1371/journal.pone.0298808. PMID: 38598488; PMCID: PMC11006186.

25. Du W, Menjivar RE, Donahue KL, Kadiyala P, Velez-Delgado A, Brown KL, Watkoske HR, He X, Carpenter ES, Angeles CV, Zhang Y, Pasca di Magliano M. WNT signaling in the tumor microenvironment promotes immunosuppression in murine pancreatic cancer. J Exp Med. 2023 Jan 2;220(1):e20220503. doi: 10.1084/jem.20220503. Epub 2022 Oct 14. PMID: 36239683; PMCID: PMC9577101.

26. Aguilera KY, Dawson DW. WNT Ligand Dependencies in Pancreatic Cancer. Front Cell Dev Biol. 2021 Apr 28;9:671022. doi: 10.3389/fcell.2021.671022. PMID: 33996827; PMCID: PMC8113755.

27. Zhong Y, Wang Z, Fu B, Pan F, Yachida S, Dhara M, Albesiano E, Li L, Naito Y, Vilardell F, Cummings C, Martinelli P, Li A, Yonescu R, Ma Q, Griffin CA, Real FX, Iacobuzio-Donahue CA. GATA6 activates Wnt signaling in pancreatic cancer by negatively regulating the Wnt antagonist Dickkopf-1. PLoS One. 2011;6(7):e22129. doi: 10.1371/journal.pone.0022129. Epub 2011 Jul 19. PMID: 21811562; PMCID: PMC3139620.

28. Cui J, Jiang W, Wang S, Wang L, Xie K. Role of Wnt/β-catenin signaling in drug resistance of pancreatic cancer. Curr Pharm Des. 2012;18(17):2464–71. doi: 10.2174/13816128112092464. PMID: 22372504.

29. Zhang Q, Meng XK, Wang WX, Zhang RM, Zhang T, Ren JJ. The Wnt/β-catenin signaling pathway mechanism for pancreatic cancer chemoresistance in a three-dimensional cancer microenvironment. Am J Transl Res. 2016 Oct 15;8(10):4490–4498. PMID: 27830034; PMCID: PMC5095343.

30. Ram Makena M, Gatla H, Verlekar D, Sukhavasi S, K Pandey M, C Pramanik K. Wnt/β-Catenin Signaling: The Culprit in Pancreatic Carcinogenesis and Therapeutic Resistance. Int J Mol Sci. 2019 Aug 30;20(17):4242. doi: 10.3390/ijms20174242. PMID: 31480221; PMCID: PMC6747343.

31. Hebrok M, Kim SK, St Jacques B, McMahon AP, Melton DA. Regulation of pancreas development by hedgehog signaling. Development. 2000 Nov;127(22):4905–13. doi: 10.1242/dev.127.22.4905. PMID: 11044404.

32. El-Zaatari M, Daignault S, Tessier A, Kelsey G, Travnikar LA, Cantu EF, Lee J, Plonka CM, Simeone DM, Anderson MA, Merchant JL. Plasma Shh levels reduced in pancreatic cancer patients. Pancreas. 2012 Oct;41(7):1019–28. doi: 10.1097/MPA.0b013e31824a0eeb. PMID: 22513293; PMCID: PMC3404255.

33. Zhou X, Liu Z, Jang F, Xiang C, Li Y, He Y. Autocrine Sonic hedgehog attenuates inflammation in cerulein-induced acute pancreatitis in mice via upregulation of IL-10. PLoS One. 2012;7(8):e44121. doi: 10.1371/journal.pone.0044121. Epub 2012 Aug 30. PMID: 22956998; PMCID: PMC3431299.

34. Wang LW, Lin H, Lu Y, Xia W, Gao J, Li ZS. Sonic hedgehog expression in a rat model of chronic pancreatitis. World J Gastroenterol. 2014 Apr 28;20(16):4712–7. doi: 10.3748/wjg.v20.i16.4712. PMID: 24782623; PMCID: PMC4000507.

35. Wang LW, Lin H, Lu Y, Xia W, Gao J, Li ZS. Sonic hedgehog expression in a rat model of chronic pancreatitis. World J Gastroenterol. 2014 Apr 28;20(16):4712–7. doi: 10.3748/wjg.v20.i16.4712. PMID: 24782623; PMCID: PMC4000507.

36. Han L, Jiang J, Xue M, Qin T, Xiao Y, Wu E, Shen X, Ma Q, Ma J. Sonic hedgehog signaling pathway promotes pancreatic cancer pain via nerve growth factor. Reg Anesth Pain Med. 2020 Feb;45(2):137–144. doi: 10.1136/rapm-2019-100991. Epub 2019 Dec 1. PMID: 31792027.

37. Yu Y, Cheng L, Yan B, Zhou C, Qian W, Xiao Y, Qin T, Cao J, Han L, Ma Q, Ma J. Overexpression of Gremlin 1 by sonic hedgehog signaling promotes pancreatic cancer progression. Int J Oncol. 2018 Dec;53(6):2445–2457. doi: 10.3892/ijo.2018.4573. Epub 2018 Sep 26. PMID: 30272371; PMCID: PMC6203161.

38. Kelleher FC, McDermott R. Aberrations and therapeutics involving the developmental pathway Hedgehog in pancreatic cancer. Vitam Horm. 2012;88:355–78. doi: 10.1016/B978-0-12-394622-5.00016-X. PMID: 22391312.

39. Leask A. Sonic advance: CCN1 regulates sonic hedgehog in pancreatic cancer. J Cell Commun Signal. 2013 Mar;7(1):61–2. doi: 10.1007/s12079-012-0187-x. Epub 2012 Dec 20. PMID: 23255052; PMCID: PMC3590359.

40. Bausch D, Fritz S, Bolm L, Wellner UF, Fernandez-Del-Castillo C, Warshaw AL, Thayer SP, Liss AS. Hedgehog signaling promotes angiogenesis directly and indirectly in pancreatic cancer. Angiogenesis. 2020 Aug;23(3):479–492. doi: 10.1007/s10456-020-09725-x. Epub 2020 May 22. PMID: 32444947.

41. Bailey JM, Swanson BJ, Hamada T, Eggers JP, Singh PK, Caffery T, Ouellette MM, Hollingsworth MA. Sonic hedgehog promotes desmoplasia in pancreatic cancer. Clin Cancer Res. 2008 Oct 1;14(19):5995–6004. doi: 10.1158/1078-0432.CCR-08-0291. PMID: 18829478; PMCID: PMC2782957.

42. Gu D, Schlotman KE, Xie J. Deciphering the role of hedgehog signaling in pancreatic cancer. J Biomed Res. 2016 Sep;30(5):353–360. doi: 10.7555/JBR.30.20150107. Epub 2016 Apr 10. PMID: 27346466; PMCID: PMC5044707.

43. Hernandez G, Luo T, Javed TA, Wen L, Kalwat MA, Vale K, Ammouri F, Husain SZ, Kliewer SA, Mangelsdorf DJ. Pancreatitis is an FGF21-deficient state that is corrected by replacement therapy. Sci Transl Med. 2020 Jan 8;12(525):eaay5186. doi: 10.1126/scitranslmed.aay5186. PMID: 31915301; PMCID: PMC7034981.

44. Tu HJ, Zhao CF, Chen ZW, Lin W, Jiang YC. Fibroblast Growth Factor (FGF) Signaling Protects Against Acute Pancreatitis-Induced Damage by Modulating Inflammatory Responses. Med Sci Monit. 2020 Apr 13;26:e920684. doi: 10.12659/MSM.920684. PMID: 32283546; PMCID: PMC7171432.

45. Kornmann M, Beger HG, Korc M. Role of fibroblast growth factors and their receptors in pancreatic cancer and chronic pancreatitis. Pancreas. 1998 Aug;17(2):169–75. doi: 10.1097/00006676-199808000-00010. PMID: 9700949.

46. Ishiwata T, Naito Z, Lu YP, Kawahara K, Fujii T, Kawamoto Y, Teduka K, Sugisaki Y. Differential distribution of fibroblast growth factor (FGF)-7 and FGF-10 in L-arginine-induced acute pancreatitis. Exp Mol Pathol. 2002 Dec;73(3):181–90. doi: 10.1006/exmp.2002.2472. PMID: 12565793.

47. Ndlovu R, Deng LC, Wu J, Li XK, Zhang JS. Fibroblast Growth Factor 10 in Pancreas Development and Pancreatic Cancer. Front Genet. 2018 Oct 29;9:482. doi: 10.3389/fgene.2018.00482. PMID: 30425728; PMCID: PMC6219204.

48. Dettmer R, Cirksena K, Münchhoff J, Kresse J, Diekmann U, Niwolik I, Buettner FFR, Naujok O. FGF2 Inhibits Early Pancreatic Lineage Specification during Differentiation of Human Embryonic Stem Cells. Cells. 2020 Aug 20;9(9):1927. doi: 10.3390/cells9091927. PMID: 32825270; PMCID: PMC7565644.

49. Carter EP, Coetzee AS, Tomas Bort E, Wang Q, Kocher HM, Grose RP. Dissecting FGF Signalling to Target Cellular Crosstalk in Pancreatic Cancer. Cells. 2021 Apr 8;10(4):847. doi: 10.3390/cells10040847. PMID: 33918004; PMCID: PMC8068358.

50. Hasegawa Y, Takada M, Yamamoto M, Saitoh Y. The gradient of basic fibroblast growth factor concentration in human pancreatic cancer cell invasion. Biochem Biophys Res Commun. 1994 May 16;200(3):1435–9. doi: 10.1006/bbrc.1994.1611. PMID: 8185597.

51. Gnatenko DA, Kopantsev EP, Sverdlov ED. Rol’ signal’nogo puti FGF/FGFR v kantserogeneze podzheludochnoĭ zhelezy [Role of fibroblast growth factors in pancreatic cancer]. Biomed Khim. 2016 Nov;62(6):622–629. Russian. doi: 10.18097/PBMC20166206622. PMID: 28026804.

52. Wagner M, Lopez ME, Cahn M, Korc M. Suppression of fibroblast growth factor receptor signaling inhibits pancreatic cancer growth in vitro and in vivo. Gastroenterology. 1998 Apr;114(4):798–807. doi: 10.1016/s0016-5085(98)70594-3. PMID: 9516401.

53. Coleman SJ, Chioni AM, Ghallab M, Anderson RK, Lemoine NR, Kocher HM, Grose RP. Nuclear translocation of FGFR1 and FGF2 in pancreatic stellate cells facilitates pancreatic cancer cell invasion. EMBO Mol Med. 2014 Apr;6(4):467–81. doi: 10.1002/emmm.201302698. Epub 2014 Feb 6. PMID: 24503018; PMCID: PMC3992074.

54. Kang X, Lin Z, Xu M, Pan J, Wang ZW. Deciphering role of FGFR signalling pathway in pancreatic cancer. Cell Prolif. 2019 May;52(3):e12605. doi: 10.1111/cpr.12605. Epub 2019 Apr 3. PMID: 30945363; PMCID: PMC6536421.

55. Slater SD, Williamson RC, Foster CS. Expression of transforming growth factor-beta 1 in chronic pancreatitis. Digestion. 1995;56(3):237–41. doi: 10.1159/000201249. PMID: 7657050.

56. Gough NR, Xiang X, Mishra L. TGF-β Signaling in Liver, Pancreas, and Gastrointestinal Diseases and Cancer. Gastroenterology. 2021 Aug;161(2):434–452.e15. doi: 10.1053/j.gastro.2021.04.064. Epub 2021 Apr 30. PMID: 33940008; PMCID: PMC8841117.

57. Truty MJ, Urrutia R. Basics of TGF-beta and pancreatic cancer. Pancreatology. 2007;7(5-6):423–35. doi: 10.1159/000108959. Epub 2007 Sep 25. PMID: 17898532.

58. Lee JH, Lee JH, Rane SG. TGF-β Signaling in Pancreatic Islet β Cell Development and Function. Endocrinology. 2021 Mar 1;162(3):bqaa233. doi: 10.1210/endocr/bqaa233. PMID: 33349851; PMCID: PMC8240135.

59. Zhou Q, Xia S, Guo F, Hu F, Wang Z, Ni Y, Wei T, Xiang H, Shang D. Transforming growth factor-β in pancreatic diseases: Mechanisms and therapeutic potential. Pharmacol Res. 2019 Apr;142:58–69. doi: 10.1016/j.phrs.2019.01.038. Epub 2019 Jan 22. PMID: 30682425.

60. Rane SG, Lee JH, Lin HM. Transforming growth factor-beta pathway: role in pancreas development and pancreatic disease. Cytokine Growth Factor Rev. 2006 Feb-Apr;17(1-2):107–19. doi: 10.1016/j.cytogfr.2005.09.003. Epub 2005 Oct 27. PMID: 16257256.

61. Hussain SM, Kansal RG, Alvarez MA, Hollingsworth TJ, Elahi A, Miranda-Carboni G, Hendrick LE, Pingili AK, Albritton LM, Dickson PV, Deneve JL, Yakoub D, Hayes DN, Kurosu M, Shibata D, Makowski L, Glazer ES. Role of TGF-β in pancreatic ductal adenocarcinoma progression and PD-L1 expression. Cell Oncol (Dordr). 2021 Jun;44(3):673–687. doi: 10.1007/s13402-021-00594-0. Epub 2021 Mar 10. PMID: 33694102.

62. Alvarez MA, Freitas JP, Mazher Hussain S, Glazer ES. TGF-β Inhibitors in Metastatic Pancreatic Ductal Adenocarcinoma. J Gastrointest Cancer. 2019 Jun;50(2):207–213. doi: 10.1007/s12029-018-00195-5. PMID: 30891677.

63. Qiang L, Hoffman MT, Ali LR, Castillo JI, Kageler L, Temesgen A, Lenehan P, Wang SJ, Bello E, Cardot-Ruffino V, Uribe GA, Yang A, Dougan M, Aguirre AJ, Raghavan S, Pelletier M, Cremasco V, Dougan SK. Transforming Growth Factor-β Blockade in Pancreatic Cancer Enhances Sensitivity to Combination Chemotherapy. Gastroenterology. 2023 Oct;165(4):874–890.e10. doi: 10.1053/j.gastro.2023.05.038. Epub 2023 May 30. PMID: 37263309; PMCID: PMC10526623.

64. Hilbig A, Oettle H. Transforming growth factor beta in pancreatic cancer. Curr Pharm Biotechnol. 2011 Dec;12(12):2158–64. doi: 10.2174/138920111798808356. PMID: 21619533.

65. Shen W, Tao GQ, Zhang Y, Cai B, Sun J, Tian ZQ. TGF-β in pancreatic cancer initiation and progression: two sides of the same coin. Cell Biosci. 2017 Aug 7;7:39. doi: 10.1186/s13578-017-0168-0. PMID: 28794854; PMCID: PMC5545849.

66. Javle M, Li Y, Tan D, Dong X, Chang P, Kar S, Li D. Biomarkers of TGF-β signaling pathway and prognosis of pancreatic cancer. PLoS One. 2014 Jan 20;9(1):e85942. doi: 10.1371/journal.pone.0085942. PMID: 24465802; PMCID: PMC3896410.

67. Principe DR, DeCant B, Mascariñas E, Wayne EA, Diaz AM, Akagi N, Hwang R, Pasche B, Dawson DW, Fang D, Bentrem DJ, Munshi HG, Jung B, Grippo PJ. TGFβ Signaling in the Pancreatic Tumor Microenvironment Promotes Fibrosis and Immune Evasion to Facilitate Tumorigenesis. Cancer Res. 2016 May 1;76(9):2525–39. doi: 10.1158/0008-5472.CAN-15-1293. Epub 2016 Mar 15. PMID: 26980767; PMCID: PMC4873388.

68. Cao Y, Drake M, Davis J, Yang B, Ko TC. Opposing Roles of BMP and TGF-β Signaling Pathways in Pancreatitis: Mechanisms and Therapeutic Implication. Adv Res Gastroenterol Hepatol. 2019;13(5):555871. Epub 2019 Sep 4. PMID: 32211568; PMCID: PMC7093075.

69. Sui L, Geens M, Sermon K, Bouwens L, Mfopou JK. Role of BMP signaling in pancreatic progenitor differentiation from human embryonic stem cells. Stem Cell Rev Rep. 2013 Oct;9(5):569–77. doi: 10.1007/s12015-013-9435-6. PMID: 23468018.

70. Klein D, Álvarez-Cubela S, Lanzoni G, Vargas N, Prabakar KR, Boulina M, Ricordi C, Inverardi L, Pastori RL, Domínguez-Bendala J. BMP-7 Induces Adult Human Pancreatic Exocrine-to-Endocrine Conversion. Diabetes. 2015 Dec;64(12):4123–34. doi: 10.2337/db15-0688. Epub 2015 Aug 25. PMID: 26307584; PMCID: PMC4657585.

71. Vukicevic S, Grgurevic L. BMP-6 and mesenchymal stem cell differentiation. Cytokine Growth Factor Rev. 2009 Oct-Dec;20(5-6):441–8. doi: 10.1016/j.cytogfr.2009.10.020. Epub 2009 Nov 8. PMID: 19900832.

72. Spagnoli FM, Brivanlou AH. The Gata5 target, TGIF2, defines the pancreatic region by modulating BMP signals within the endoderm. Development. 2008 Feb;135(3):451–61. doi: 10.1242/dev.008458. Epub 2007 Dec 19. PMID: 18094028.

73. Chmielowiec J, Szlachcic WJ, Yang D, Scavuzzo MA, Wamble K, Sarrion-Perdigones A, Sabek OM, Venken KJT, Borowiak M. Human pancreatic microenvironment promotes β-cell differentiation via non-canonical WNT5A/JNK and BMP signaling. Nat Commun. 2022 Apr 12;13(1):1952. doi: 10.1038/s41467-022-29646-1. PMID: 35414140; PMCID: PMC9005503.

74. Gordon KJ, Kirkbride KC, How T, Blobe GC. Bone morphogenetic proteins induce pancreatic cancer cell invasiveness through a Smad1-dependent mechanism that involves matrix metalloproteinase-2. Carcinogenesis. 2009 Feb;30(2):238–48. doi: 10.1093/carcin/bgn274. Epub 2008 Dec 4. PMID: 19056927; PMCID: PMC2639045.

75. Kleeff J, Maruyama H, Ishiwata T, Sawhney H, Friess H, Büchler MW, Korc M. Bone morphogenetic protein 2 exerts diverse effects on cell growth in vitro and is expressed in human pancreatic cancer in vivo. Gastroenterology. 1999 May;116(5):1202–16. doi: 10.1016/s0016-5085(99)70024-7. PMID: 10220513.

76. Gordon KJ, Kirkbride KC, How T, Blobe GC. Bone morphogenetic proteins induce pancreatic cancer cell invasiveness through a Smad1-dependent mechanism that involves matrix metalloproteinase-2. Carcinogenesis. 2009 Feb;30(2):238–48. doi: 10.1093/carcin/bgn274. Epub 2008 Dec 4. PMID: 19056927; PMCID: PMC2639045.

77. Mines D, Gu Y, Kou TD, Cooper GS. Recombinant human bone morphogenetic protein-2 and pancreatic cancer: a retrospective cohort study. Pharmacoepidemiol Drug Saf. 2011 Feb;20(2):111–8. doi: 10.1002/pds.2057. Epub 2010 Dec 9. PMID: 21254281.

78. Virtanen S, Alarmo EL, Sandström S, Ampuja M, Kallioniemi A. Bone morphogenetic protein-4 and -5 in pancreatic cancer--novel bidirectional players. Exp Cell Res. 2011 Sep 10;317(15):2136–46. doi: 10.1016/j.yexcr.2011.06.001. Epub 2011 Jun 17. PMID: 21704030.

79. Lan L, Evan T, Li H, Hussain A, Ruiz EJ, Zaw Thin M, Ferreira RMM, Ps H, Riising EM, Zen Y, Almagro J, Ng KW, Soro-Barrio P, Nelson J, Koifman G, Carvalho J, Nye EL, He Y, Zhang C, Sadanandam A, Behrens A. GREM1 is required to maintain cellular heterogeneity in pancreatic cancer. Nature. 2022 Jul;607(7917):163–168. doi: 10.1038/s41586-022-04888-7. Epub 2022 Jun 29. PMID: 35768509.

80. Cheng Z, Cui W, Ding Y, Liu T, Liu W, Qin Y, Xia W, Xu J, Zhang Y, Zou X. BMP8B mediates the survival of pancreatic cancer cells and regulates the progression of pancreatic cancer. Oncol Rep. 2014 Nov;32(5):1861–6. doi: 10.3892/or.2014.3413. Epub 2014 Aug 18. PMID: 25176058.

81. Stanciu S, Ionita-Radu F, Stefani C, Miricescu D, Stanescu-Spinu II, Greabu M, Ripszky Totan A, Jinga M. Targeting PI3K/AKT/mTOR Signaling Pathway in Pancreatic Cancer: From Molecular to Clinical Aspects. Int J Mol Sci. 2022 Sep 4;23(17):10132. doi: 10.3390/ijms231710132. PMID: 36077529; PMCID: PMC9456549.

82. Mortazavi M, Moosavi F, Martini M, Giovannetti E, Firuzi O. Prospects of targeting PI3K/AKT/mTOR pathway in pancreatic cancer. Crit Rev Oncol Hematol. 2022 Aug;176:103749. doi: 10.1016/j.critrevonc.2022.103749. Epub 2022 Jun 18. PMID: 35728737.

83. Wang F, Yan X, Hua Y, Song J, Liu D, Yang C, Peng F, Kang F, Hui Y. PI3K/AKT/mTOR pathway and its related molecules participate in PROK1 silence-induced anti-tumor effects on pancreatic cancer. Open Life Sci. 2023 Apr 10;18(1):20220538. doi: 10.1515/biol-2022-0538. PMID: 37070074; PMCID: PMC10105552.

84. Stanciu S, Ionita-Radu F, Stefani C, Miricescu D, Stanescu-Spinu II, Greabu M, Ripszky Totan A, Jinga M. Targeting PI3K/AKT/mTOR Signaling Pathway in Pancreatic Cancer: From Molecular to Clinical Aspects. Int J Mol Sci. 2022 Sep 4;23(17):10132. doi: 10.3390/ijms231710132. PMID: 36077529; PMCID: PMC9456549.

85. Thibault B, Ramos-Delgado F, Pons-Tostivint E, Therville N, Cintas C, Arcucci S, Cassant-Sourdy S, Reyes-Castellanos G, Tosolini M, Villard AV, Cayron C, Baer R, Bertrand-Michel J, Pagan D, Ferreira Da Mota D, Yan H, Falcomatà C, Muscari F, Bournet B, Delord JP, Aksoy E, Carrier A, Cordelier P, Saur D, Basset C, Guillermet-Guibert J. Pancreatic cancer intrinsic PI3Kα activity accelerates metastasis and rewires macrophage component. EMBO Mol Med. 2021 Jul 7;13(7):e13502. doi: 10.15252/emmm.202013502. Epub 2021 May 25. PMID: 34033220; PMCID: PMC8261517.

86. Faleiro I, Roberto VP, Demirkol Canli S, Fraunhoffer NA, Iovanna J, Gure AO, Link W, Castelo-Branco P. DNA Methylation of PI3K/AKT Pathway-Related Genes Predicts Outcome in Patients with Pancreatic Cancer: A Comprehensive Bioinformatics-Based Study. Cancers (Basel). 2021 Dec 17;13(24):6354. doi: 10.3390/cancers13246354. PMID: 34944974; PMCID: PMC8699150.

87. Mehra S, Deshpande N, Nagathihalli N. Targeting PI3K Pathway in Pancreatic Ductal Adenocarcinoma: Rationale and Progress. Cancers (Basel). 2021 Sep 2;13(17):4434. doi: 10.3390/cancers13174434. PMID: 34503244; PMCID: PMC8430624.

88. Ebrahimi S, Hosseini M, Shahidsales S, Maftouh M, Ferns GA, Ghayour-Mobarhan M, Hassanian SM, Avan A. Targeting the Akt/PI3K Signaling Pathway as a Potential Therapeutic Strategy for the Treatment of Pancreatic Cancer. Curr Med Chem. 2017;24(13):1321–1331. doi: 10.2174/0929867324666170206142658. PMID: 28176634.

89. Baer R, Cintas C, Therville N, Guillermet-Guibert J. Implication of PI3K/Akt pathway in pancreatic cancer: When PI3K isoforms matter? Adv Biol Regul. 2015 Sep;59:19–35. doi: 10.1016/j.jbior.2015.05.001. Epub 2015 Jun 20. PMID: 26166735.

90. Xie P, Tan SY, Li HF, Tang HD, Zhou JH. Transcriptome data-based status of PI3K/AKT/mTOR pathway indicates heterogeneity and immune modulation in patients with pancreatic ductal adenocarcinoma. J Gene Med. 2024 Jan;26(1):e3570. doi: 10.1002/jgm.3570. Epub 2023 Jul 22. PMID: 37482968.

91. Khan KH, Yap TA, Yan L, Cunningham D. Targeting the PI3K-AKT-mTOR signaling network in cancer. Chin J Cancer. 2013 May;32(5):253–65. doi: 10.5732/cjc.013.10057. PMID: 23642907; PMCID: PMC3845556.

92. Yu JH, Kim H. Role of janus kinase/signal transducers and activators of transcription in the pathogenesis of pancreatitis and pancreatic cancer. Gut Liver. 2012 Oct;6(4):417–22. doi: 10.5009/gnl.2012.6.4.417. Epub 2012 Aug 7. PMID: 23170143; PMCID: PMC3493719.

93. Komar HM, Serpa G, Kerscher C, Schwoegl E, Mace TA, Jin M, Yang MC, Chen CS, Bloomston M, Ostrowski MC, Hart PA, Conwell DL, Lesinski GB. Inhibition of Jak/STAT signaling reduces the activation of pancreatic stellate cells in vitro and limits caerulein-induced chronic pancreatitis in vivo. Sci Rep. 2017 May 11;7(1):1787. doi: 10.1038/s41598-017-01973-0. PMID: 28496202; PMCID: PMC5431930.

94. Gallmeier E, Schäfer C, Moubarak P, Tietz A, Plössl I, Huss R, Göke B, Wagner AC. JAK and STAT proteins are expressed and activated by IFN-gamma in rat pancreatic acinar cells. J Cell Physiol. 2005 Apr;203(1):209–16. doi: 10.1002/jcp.20216. PMID: 15493010.

95. Doi T, Ishikawa T, Okayama T, Oka K, Mizushima K, Yasuda T, Sakamoto N, Katada K, Kamada K, Uchiyama K, Handa O, Takagi T, Naito Y, Itoh Y. The JAK/STAT pathway is involved in the upregulation of PD-L1 expression in pancreatic cancer cell lines. Oncol Rep. 2017 Mar;37(3):1545–1554. doi: 10.3892/or.2017.5399. Epub 2017 Jan 23. PMID: 28112370.

96. Qiu Z, Xu F, Wang Z, Yang P, Bu Z, Cheng F, Jiang H, Li L, Zhang F. Blockade of JAK2 signaling produces immunomodulatory effect to preserve pancreatic homeostasis in severe acute pancreatitis. Biochem Biophys Rep. 2021 Sep 17;28:101133. doi: 10.1016/j.bbrep.2021.101133. PMID: 34584986; PMCID: PMC8453217.

97. Biffi G, Oni TE, Spielman B, Hao Y, Elyada E, Park Y, Preall J, Tuveson DA. IL1-Induced JAK/STAT Signaling Is Antagonized by TGFβ to Shape CAF Heterogeneity in Pancreatic Ductal Adenocarcinoma. Cancer Discov. 2019 Feb;9(2):282–301. doi: 10.1158/2159-8290.CD-18-0710. Epub 2018 Oct 26. PMID: 30366930; PMCID: PMC6368881.

98. Lin XM, Chen H, Zhan XL. MiR-203 regulates JAK-STAT pathway in affecting pancreatic cancer cells proliferation and apoptosis by targeting SOCS3. Eur Rev Med Pharmacol Sci. 2019 Aug;23(16):6906–6913. doi: 10.26355/eurrev_201908_18730. PMID: 31486490.

99. Shrestha H, Rädler PD, Dennaoui R, Wicker MN, Rajbhandari N, Sun Y, Peck AR, Vistisen K, Triplett AA, Beydoun R, Sterneck E, Saur D, Rui H, Wagner KU. The Janus kinase 1 is critical for pancreatic cancer initiation and progression. Cell Rep. 2024 May 28;43(5):114202. doi: 10.1016/j.celrep.2024.114202. Epub 2024 May 10. PMID: 38733583; PMCID: PMC11194014.

100. Yu JH, Kim H. Role of janus kinase/signal transducers and activators of transcription in the pathogenesis of pancreatitis and pancreatic cancer. Gut Liver. 2012 Oct;6(4):417–22. doi: 10.5009/gnl.2012.6.4.417. Epub 2012 Aug 7. PMID: 23170143; PMCID: PMC3493719.

101. Raymant M, Astuti Y, Alvaro-Espinosa L, Green D, Quaranta V, Bellomo G, Glenn M, Chandran-Gorner V, Palmer DH, Halloran C, Ghaneh P, Henderson NC, Morton JP, Valiente M, Mielgo A, Schmid MC. Macrophage-fibroblast JAK/STAT dependent crosstalk promotes liver metastatic outgrowth in pancreatic cancer. Nat Commun. 2024 Apr 27;15(1):3593. doi: 10.1038/s41467-024-47949-3. PMID: 38678021; PMCID: PMC11055860.

102. Lu C, Talukder A, Savage NM, Singh N, Liu K. JAK-STAT-mediated chronic inflammation impairs cytotoxic T lymphocyte activation to decrease anti-PD-1 immunotherapy efficacy in pancreatic cancer. Oncoimmunology. 2017 Feb 10;6(3):e1291106. doi: 10.1080/2162402X.2017.1291106. PMID: 28405527; PMCID: PMC5384417.

103. Pang C, Gu Y, Ding Y, Ma C, Yv W, Wang Q, Meng B. Several genes involved in the JAK-STAT pathway may act as prognostic markers in pancreatic cancer identified by microarray data analysis. Medicine (Baltimore). 2018 Dec;97(50):e13297. doi: 10.1097/MD.0000000000013297. PMID: 30557977; PMCID: PMC6320066.

104. Song Y, Tang MY, Chen W, Wang Z, Wang SL. High JAK2 Protein Expression Predicts a Poor Prognosis in Patients with Resectable Pancreatic Ductal Adenocarcinoma. Dis Markers. 2020 Sep 21;2020:7656031. doi: 10.1155/2020/7656031. PMID: 33029256; PMCID: PMC7528024.

105. Lu C, Talukder A, Savage NM, Singh N, Liu K. JAK-STAT-mediated chronic inflammation impairs cytotoxic T lymphocyte activation to decrease anti-PD-1 immunotherapy efficacy in pancreatic cancer. Oncoimmunology. 2017 Feb 10;6(3):e1291106. doi: 10.1080/2162402X.2017.1291106. PMID: 28405527; PMCID: PMC5384417.

106. Ardestani A, Maedler K. The Hippo Signaling Pathway in Pancreatic β-Cells: Functions and Regulations. Endocr Rev. 2018 Feb 1;39(1):21–35. doi: 10.1210/er.2017-00167. PMID: 29053790.

107. George NM, Day CE, Boerner BP, Johnson RL, Sarvetnick NE. Hippo signaling regulates pancreas development through inactivation of Yap. Mol Cell Biol. 2012 Dec;32(24):5116–28. doi: 10.1128/MCB.01034-12. Epub 2012 Oct 15. PMID: 23071096; PMCID: PMC3510525.

108. Wu Y, Aegerter P, Nipper M, Ramjit L, Liu J, Wang P. Hippo Signaling Pathway in Pancreas Development. Front Cell Dev Biol. 2021 May 17;9:663906. doi: 10.3389/fcell.2021.663906. PMID: 34079799; PMCID: PMC8165189.

109. Ansari D, Ohlsson H, Althini C, Bauden M, Zhou Q, Hu D, Andersson R. The Hippo Signaling Pathway in Pancreatic Cancer. Anticancer Res. 2019 Jul;39(7):3317–3321. doi: 10.21873/anticanres.13474. PMID: 31262852.

110. Tamura T, Kodama T, Sato K, Murai K, Yoshioka T, Shigekawa M, Yamada R, Hikita H, Sakamori R, Akita H, Eguchi H, Johnson RL, Yokoi H, Mukoyama M, Tatsumi T, Takehara T. Dysregulation of PI3K and Hippo signaling pathways synergistically induces chronic pancreatitis via CTGF upregulation. J Clin Invest. 2021 Jul 1;131(13):e143414. doi: 10.1172/JCI143414. PMID: 34032634; PMCID: PMC8245178.

111. Gao T, Zhou D, Yang C, Singh T, Penzo-Méndez A, Maddipati R, Tzatsos A, Bardeesy N, Avruch J, Stanger BZ. Hippo signaling regulates differentiation and maintenance in the exocrine pancreas. Gastroenterology. 2013 Jun;144(7):1543–53, 1553.e1. doi: 10.1053/j.gastro.2013.02.037. Epub 2013 Feb 27. PMID: 23454691; PMCID: PMC3665616.

112. Ansari D, Ohlsson H, Althini C, Bauden M, Zhou Q, Hu D, Andersson R. The Hippo Signaling Pathway in Pancreatic Cancer. Anticancer Res. 2019 Jul;39(7):3317–3321. doi: 10.21873/anticanres.13474. PMID: 31262852.

113. Drexler R, Küchler M, Wagner KC, Reese T, Feyerabend B, Kleine M, Oldhafer KJ. The clinical relevance of the Hippo pathway in pancreatic ductal adenocarcinoma. J Cancer Res Clin Oncol. 2021 Feb;147(2):373–391. doi: 10.1007/s00432-020-03427-z. Epub 2020 Oct 24. PMID: 33098447; PMCID: PMC7817599.

114. Su W, Zhu S, Chen K, Yang H, Tian M, Fu Q, Shi G, Feng S, Ren D, Jin X, Yang C. Overexpressed WDR3 induces the activation of Hippo pathway by interacting with GATA4 in pancreatic cancer. J Exp Clin Cancer Res. 2021 Mar 1;40(1):88. doi: 10.1186/s13046-021-01879-w. PMID: 33648545; PMCID: PMC7923337.

115. Ren D, Sun Y, Zhang D, Li D, Liu Z, Jin X, Wu H. SGLT2 promotes pancreatic cancer progression by activating the Hippo signaling pathway via the hnRNPK-YAP1 axis. Cancer Lett. 2021 Oct 28;519:277–288. doi: 10.1016/j.canlet.2021.07.035. Epub 2021 Jul 24. PMID: 34314754.

116. Lv L, Zhou X. Targeting Hippo signaling in cancer: novel perspectives and therapeutic potential. MedComm (2020). 2023 Oct 3;4(5):e375. doi: 10.1002/mco2.375. PMID: 37799806; PMCID: PMC10547939.

